# A highly divergent cryptic SARS-CoV-2 lineage exhibits strong receptor binding and immune evasion

**DOI:** 10.1101/2025.11.18.25340485

**Authors:** Ziqi Feng, Qi Wen Teo, Barikisu A. Ibrahim, Wenkan Liu, Ding Zhang, Kevin J. Mao, Evan K. Shao, Devon A. Gregory, Ian A. Wilson, Marc C. Johnson, Meng Yuan, Nicholas C. Wu

**Author notes:** These authors contributed equally (Ziqi Feng, Qi Wen Teo). Correspondence (M.C.J.), (M.Y.), (N.C.W.).

## Abstract

SARS-CoV-2 genomic surveillance in wastewater enables the identification of cryptic lineages, often with numerous mutations not observed in circulating variants. Here, we discovered a unique cryptic SARS-CoV-2 lineage (designated NJ) from New Jersey wastewater that carries many rare mutations and exhibits the highest binding affinity to the hACE2 receptor among all tested SARS-CoV-2 variant RBDs, including the latest Omicron lineages. Moreover, NJ shows extensive evasion against all five major classes of neutralizing antibodies to SARS-CoV-2. Deep mutational scanning further demonstrates the involvement of epistasis in the evolution of the NJ-RBD. Crystal structures of NJ-RBD in complex with hACE2 and with a broadly neutralizing antibody CC25.4 reveal key amino acid substitutions that affect receptor binding, antibody evasion, and explain its unique molecular characteristics. Overall, these findings not only elucidate the structural and functional features of a cryptic SARS-CoV-2 lineage, but also provide insights into potential future evolution of SARS-CoV-2.

## Main

Over the past five years, severe acute respiratory syndrome coronavirus 2 (SARS-CoV-2) has continued to circulate globally and mutate, causing immense harm to public health and the economy. Wastewater viral sequencing has developed into a cost-effective surveillance method that tracks the dynamics of virus evolution at the population level within specific geographical areas^1–4^. Compared with conventional clinical sampling, wastewater viral sequencing can detect mutation information earlier and help predict the evolutionary trajectory of the virus^5^. Moreover, wastewater viral sequencing captures data from all infected individuals, including those with mild symptoms who did not seek medical attention or are asymptomatic, thereby complementing other public health surveillance data^6,7^.

With advancements in sequencing technology and analytical tools, it is now possible to identify rare variants in wastewater samples with confidence^8^. SARS-CoV-2 strains containing mutations that differ substantially from those in any variants of concern (VOCs) and variants of interest (VOIs) are referred to as cryptic lineages^6,7,9^. These lineages often originate from individuals with chronic SARS-CoV-2 infections, in which extensive within-host virus evolution has occurred independently of circulating strains^6,9^. Interestingly, some unique spike amino acid substitutions in previously discovered cryptic SARS-CoV-2 lineages have since been found in VOCs/VOIs, reflecting evolutionary convergence^7,9^. For example, during the early stages of the COVID-19 pandemic (pre-Omicron), wastewater viral sequencing identified some substitutions in cryptic lineages that later emerged in the Omicron BA.1 spike protein, such as N440K, G446S, S477N, T478K, E484A, Q493R, G496S, Q498R, N501Y, and Y505H^7^. Some of these substitutions, primarily located in the receptor binding domain (RBD), enhanced binding to the receptor human angiotensin converting enzyme 2 (hACE2) or enabled evasion from neutralizing antibodies. These observations highlight the importance and foresight of wastewater viral sequencing. By contrast, many other substitutions that emerged in the cryptic SARS-CoV-2 lineages are distinct from the commonly emerged substitutions in VOCs/VOIs^7,10–13^. It is therefore important to investigate the functional consequences of the evolutionary trajectories of cryptic SARS-CoV-2 lineages.

In this study, we identified a cryptic SARS-CoV-2 lineage (designated NJ) in New Jersey wastewater containing a heavily mutated RBD distinct from any known VOC/VOI, including Omicron subvariants. Furthermore, we discovered that the NJ-RBD exhibited the highest binding affinity to hACE2 among tested SARS-CoV-2 variants, and dramatically reduced binding to all five major classes of neutralizing antibodies (classes I-V)^14,15^. The underlying mechanisms were further examined by X-ray crystallography. Additionally, deep mutational scanning demonstrated that NJ-RBD exhibited unique evolutionary potential compared to other VOCs/VOIs due to epistasis. Overall, these findings deepen our understanding of rare cryptic lineages that carry unique substitutions and have important implications in forecasting SARS-CoV-2 evolution.

## Results

### Isolation and characterization of NJ cryptic lineage

Beginning in November 2023, SARS-CoV-2 wastewater sequences uploaded to the Sequence Read Archive (SRA)^16^ were screened for evidence of cryptic SARS-CoV-2 lineages. One of the screening criteria was identifying samples that contained an exact match to the sequence TACGATCGAGGGTACAGTG. This sequence was from an RNA element at the 3’ end of the SARS-CoV-2 genome, which was deleted from all contemporary strains. It also contained the nucleotide mutation T29758G, a key indicator for identifying cryptic SARS-CoV-2 lineages^13^. As described previously, T29758G represents a reversion to the sequences found in SARS-CoV-2-related enteric bat Sarbecovirus, consistent with the notion that cryptic SARS-CoV-2 lineages predominantly replicate in the gastrointestinal tract rather than in the respiratory system^13^. This sequence was detected in a total of five samples from a New Jersey sewershed (population of 1,500,000) that were collected between November 2023 and June 2024 as part of the CDC national wastewater surveillance system (NWSS). On many collection dates, coverage of the Spike gene was limited; however, on two dates (November 20 and 22, 2023), an outlier spike sequence was observed that was nearly identical across both dates. The sequences from these two dates were analyzed to generate a consensus spike sequence, designated NJ. A schematic overview of these procedures is provided in **Fig. 1a**.

**Fig. 1:**
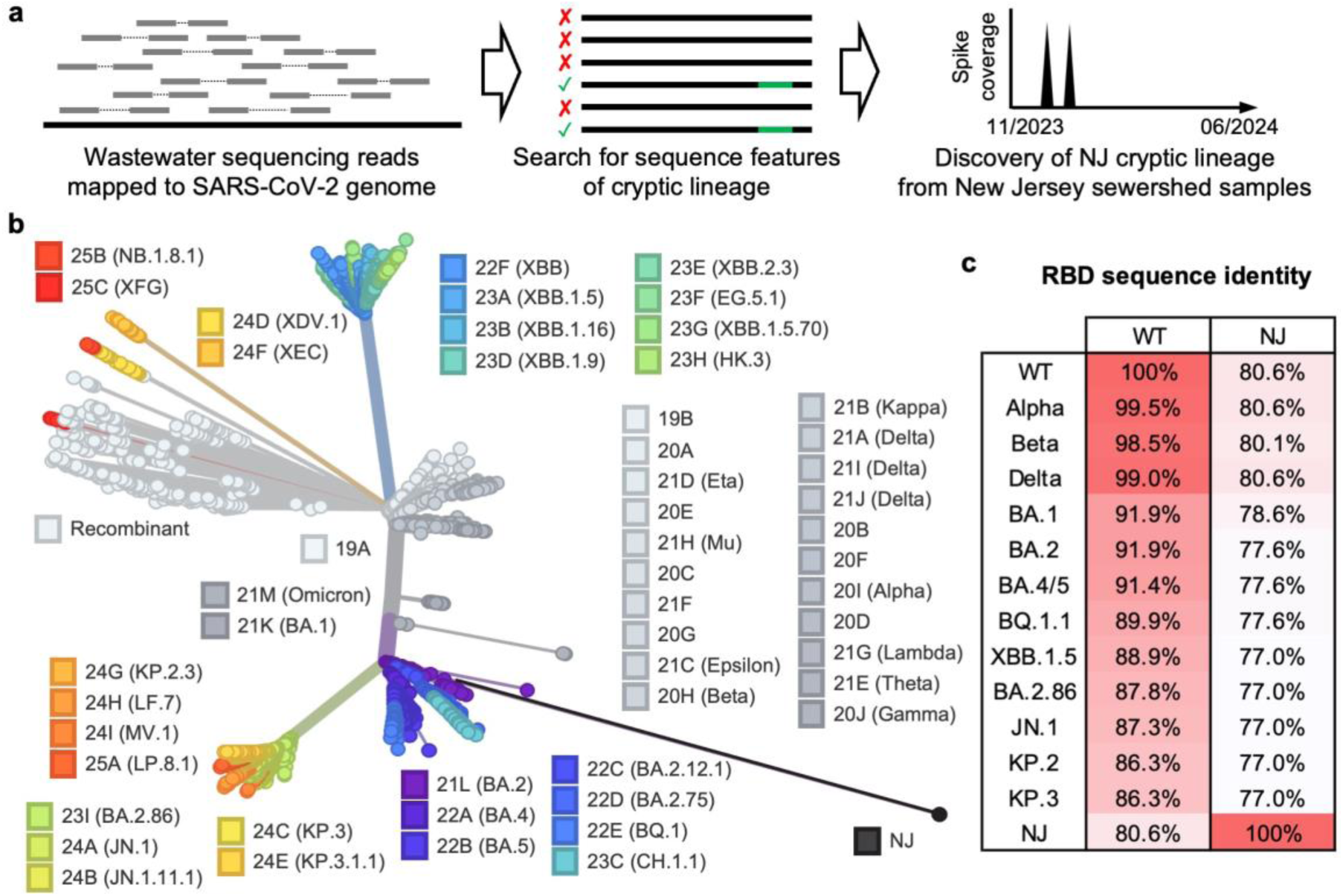
Discovery and phylogenetic analysis of the NJ cryptic lineage. **a**, Schematic workflow of the discovery of cryptic SARS-CoV-2 lineages from wastewater. **b**, Phylogenetic tree of spike gene generated by Nextclade^43^. The consensus sequences of NJ spike were uploaded onto Nextclade and compared against SARS-CoV-2 VOC/VOI spikes. NJ greatly diverges from all VOC/VOI lineages. **c**, Percentages of amino acid sequence identity between NJ and VOC/VOI RBDs.

The NJ cryptic lineage was most likely derived from a B.1 lineage that predated B.1.1.529 (Omicron), since it contained the ancestral stem-loop two motif sequence, which has been mutated in all circulating lineages since late 2022. Moreover, NJ lacked the B.1.1-defining mutations G28881A, G28882A, and G28883C. NJ also did not appear to be derived from B.1.617.2 (Delta), which was the last non-B.1.1-derived circulating lineage, since its stem-loop two motif sequence did not contain the Delta-defining mutation G29742T. Collectively, this information suggests the NJ was derived from a B.1 lineage that circulated in 2021 or earlier.

NJ-RBD had 39 amino acid substitutions compared to the wild-type RBD (WT, Wuhan-Hu-1), among which only four substitutions were observed in VOCs/VOIs, namely R403K, S477N, V483del, and Y505H (**Extended Data Fig. 1**). Interestingly, 13 of the remaining 35 substitutions have previously been observed in cryptic SARS-CoV-2 lineages identified through wastewater viral sequencing^7^. This similarity in evolutionary pathways suggest that cryptic SARS-CoV-2 lineages from different regions and time periods may encounter similar evolutionary pressures. Phylogenetic analysis demonstrated that the NJ greatly diverges from SARS-CoV-2 WT and all VOCs/VOIs (**Fig. 1b**). As SARS-CoV-2 evolved, amino acid substitutions increasingly accumulated in the RBD. For example, Alpha had 99.5% sequence identity to WT, while KP.3 had only 86.3%. Nevertheless, NJ-RBD exhibited the lowest sequence identity as compared to all known VOCs/VOIs, at just 80.6% sequence identity to WT. Additionally, NJ-RBD showed low sequence identity to other VOCs/VOIs, ranging from 77.0% to 80.6% (**Fig. 1c**).

### NJ cryptic lineage exhibits exceptionally strong binding affinity to hACE2

Previously, we and others have observed that hACE2-binding affinity has increased during the natural evolution of SARS-CoV-2^10,17^. Here, we compared the hACE2-binding affinity of NJ-RBD to that of other VOCs/VOIs using surface plasmon resonance (SPR). Remarkably, NJ-RBD exhibited the strongest binding affinity (K_D_ = 1.2 nM) among all tested VOCs/VOIs, which was 11.4-fold stronger than that of WT-RBD (K_D_ = 13.7 nM) (**Fig. 2a and Extended Data Fig. 2**). NJ-RBD displayed an association rate comparable to WT-RBD but had the lowest dissociation rate among all VOCs/VOIs (**Fig. 2b**). The low dissociation rate indicates that many unique substitutions in NJ-RBD strengthen its interaction with hACE2.

**Fig. 2:**
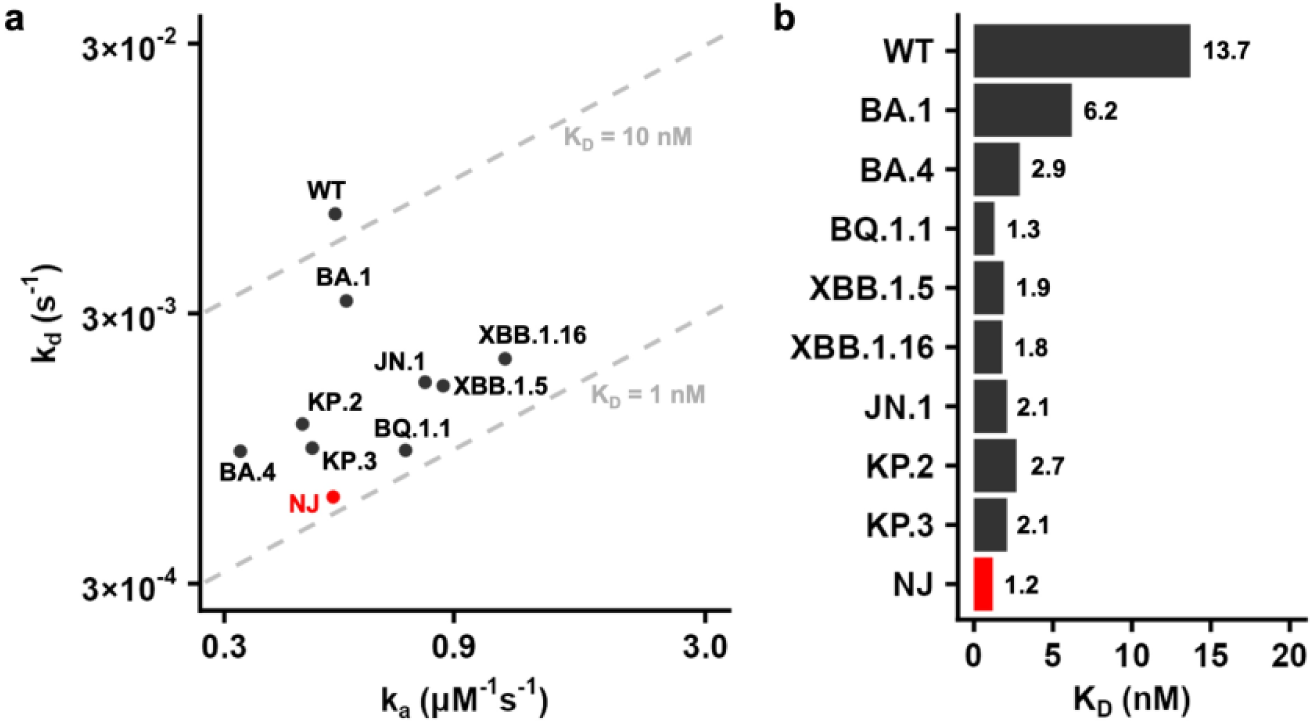
Binding affinity of NJ and VOCs/VOIs RBDs against hACE2 using SPR. **a**, Summary of binding affinity between VOC/VOI RBDs and hACE2. The association rate k_a_ (µM^-1^ s^-1^) and dissociation rate k_d_ (s^-1^) are plotted as the x-axis and y-axis, respectively. The gray dashed lines labeled 1 nM and 10 nM represent K_D_ of 1 nM and 10 nM, respectively. **b**, Binding affinity (K_D_) of the indicated RBDs to hACE2.

### Structural basis for enhanced hACE2 binding by NJ-RBD substitutions

To investigate how the unique substitutions in the NJ-RBD affect hACE2 binding, we determined the crystal structure of NJ-RBD in complex with hACE2 to 2.5 Å resolution (**Fig. 3a and Extended Data Table 1**). Structural comparison with the WT-RBD/hACE2 complex (PDB 6M0J) revealed nearly identical binding angles and positions with a Cα root mean square deviation (RMSD) value of 0.7 Å^18^ (**Extended Data Fig. 3**). Similarly, structural superimposition with other VOC/VOI RBD/hACE2 yielded Cα RMSD values of 0.6-1.2 Å. The interface area between hACE2 and RBD was also comparable (NJ-RBD/hACE2: 827 Å^2^; WT-RBD/hACE2: 843 Å^2^). However, NJ-RBD exhibited three loop shifts (residues 368-372, 445-449, and 481-484) compared to WT and other VOC/VOI RBDs^17–23^ (**Extended Data Fig. 3**). The deviations in loops 368-372 and 481-484 were primarily due to the deletion of amino acid residues 372 and 483 in NJ-RBD, while the deviation in loop 445-449 resulted from multiple substitutions (**Extended Data Fig. 3**). Despite having an almost identical footprint on hACE2 (**Extended Data Fig. 3**), the receptor-binding motif (RBM) sequences of NJ-RBD and WT-RBD were quite different, where among 25 residues directly involved in hACE2 binding, only 10 (i.e. 40%) were conserved (**Fig. 3b**).

**Fig. 3:**
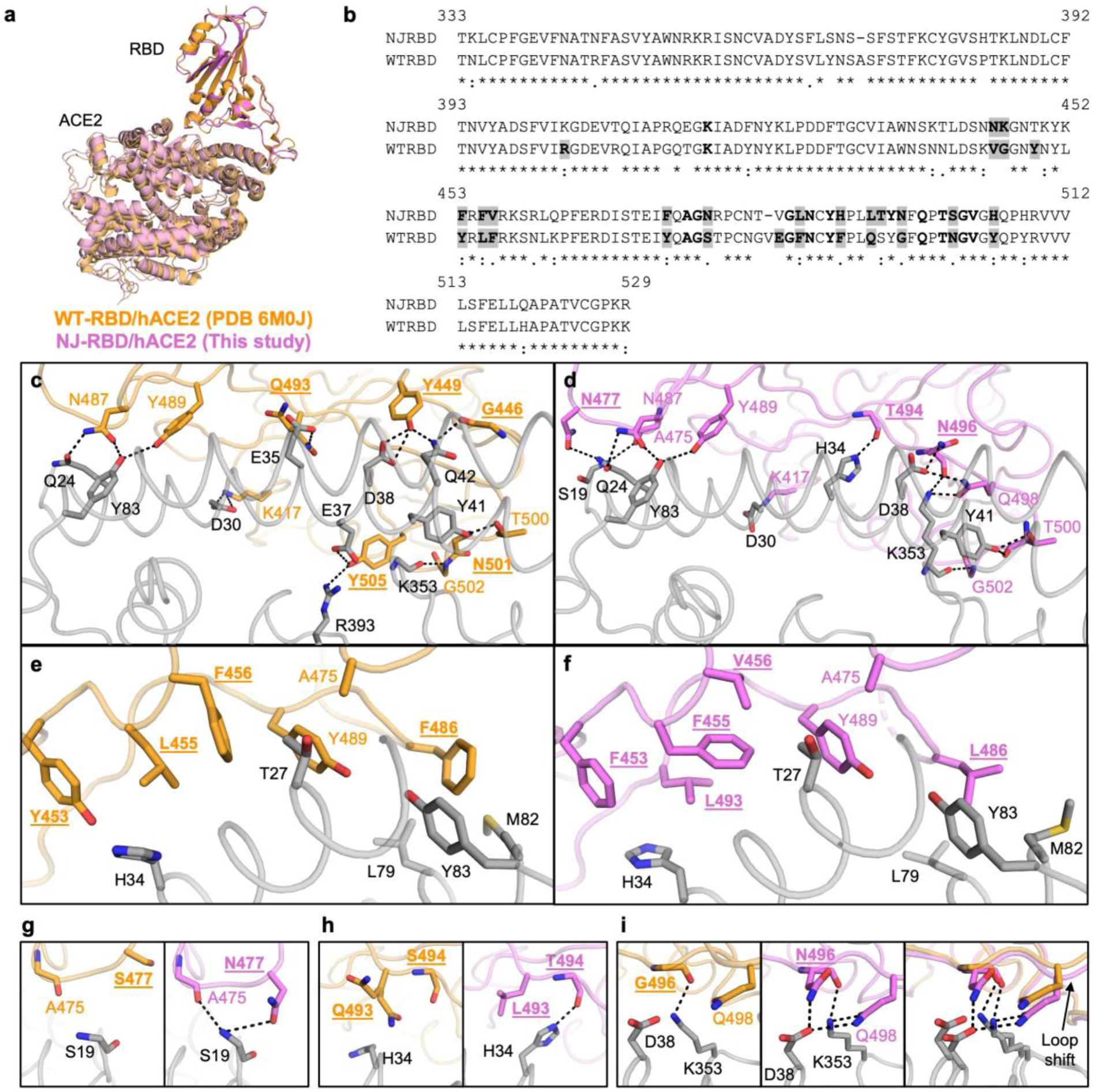
Structural comparison between NJ-RBD and WT-RBD in complex with hACE2. **a**, NJ-RBD/hACE2 (this study, pink) and WT-RBD/hACE2 (PDB 6M0J, orange)^18^ were aligned using their RBDs. **b**, Sequence alignment of NJ-RBD and WT-RBD. Residues binding to hACE2 (BSA > 0 Å^2^ as calculated by PISA^54^) are in bold. Non-conserved hACE2-binding residues are highlighted in grey. **c**,**d**, Molecular details of hydrophilic interactions (**c**) between WT-RBD and hACE2, and (**d**) between NJ-RBD with hACE2. Residues are numbered based on their positions on the SARS-CoV-2 WT spike protein sequence. H-bonds and salt bridges are represented by black dashed lines. **e**,**f**, Hydrophobic interactions (**e**) between WT-RBD and hACE2, and (**f**) between NJ-RBD with hACE2. **g**, Zoomed-in view of comparison between the N477 in NJ-RBD and its counterpart S477 in WT-RBD. **h**, Zoomed-in view of comparison between the L493 and T494 in NJ-RBD and its counterpart Q493 and S494 in WT-RBD. The two side chains of WT-Q493 represent alternate conformers of the residue. **i**, Zoomed-in view of comparison between N496 in NJ-RBD and its counterpart G496 in WT-RBD. (**c-i**) Non-conserved receptor-binding residues are in bold and underlined.

Similar to WT-RBD, NJ-RBD also formed extensive hydrophilic interactions with hACE2. These included several conserved interactions, such as the salt bridge between RBD-K417 and hACE2-D30; the H-bonds among RBD-N487, RBD-Y489, hACE2-Q24, and hACE2-Y83; the H-bond between RBD-T500 with hACE2-Y41; and the H-bond between RBD-G502 main-chain and hACE2-K353 main-chain (**Fig. 3c,d**). Furthermore, several aromatic and aliphatic receptor-binding residues in NJ-RBD formed hydrophobic interactions with hACE2. While the spatial positions of these aromatic and aliphatic residues were similar to those in WT-RBD, some of their interactions with hACE2 differed due to substitutions at residues 453, 455, 456, 486, and 493, whereas two were conserved at residues 475 and 489. (**Fig. 3e,f**).

By comparing the molecular interactions between WT-RBD and NJ-RBD with hACE2, we identified three substitutions, S477N, Q493L, and G496N, that could explain the stronger hACE2-binding affinity of NJ-RBD (**Fig. 3b**). As shown in previous studies, S477N, which was present in Omicron BA.1 and subsequent lineages, enhanced binding to hACE2 (**Extended Data Fig. 4**)^24,25^. The S477N substitution in NJ-RBD introduced an H-bond with the S19 main-chain of hACE2, and the loop shift caused the A475 main-chain to H-bond with the S19 main-chain (**Fig. 3g**). Q493L only appeared in 2024 with very low frequency (2%) (**Extended Data Fig. 4**). The Q493L substitution in the NJ cryptic lineage altered the side-chain direction of hACE2-H34, enabling L493 to form hydrophobic interactions with H34, and the T494 main-chain to H-bond with H34 due to a side-chain rotamer change (**Fig. 3h**). G496N was not detected in previous variants (**Extended Data Fig. 4**). The G496N substitution introduced an H-bond with D38 of hACE2, while the loop shift brought Q498 closer to form H-bonds with D38 and K353 (**Fig. 3i**). These substitutions facilitated additional interactions between NJ-RBD and hACE2, explaining its strongest binding affinity to date (**Fig. 2**). Together, our structural analysis substantiated the huge evolutionary potential of the RBM.

### Extensive antibody evasion by the NJ cryptic lineage

Given the number of amino acid substitutions in the NJ-RBD relative to the WT-RBD, we postulated that NJ-RBD would exhibit extensive antibody evasion. We used bio-layer interferometry (BLI) to test the binding affinities of NJ-RBD and WT-RBD to representative antibodies from the five major classes^26^ (**Fig. 4a**). Class I antibodies (e.g. CC12.3) target the receptor-binding motif (RBM), competing with the cell receptor hACE2 to achieve neutralization^27^. While WT-RBD exhibited strong binding to CC12.3 (K_D_ = 24.4 nM), NJ-RBD had no detectable binding (K_D_ >1000 nM). Only 50% (15 out of 30) of the CC12.3 epitope residues were conserved between NJ-RBD and WT-RBD (**Extended Data Fig. 5a,f**). Class II antibodies (e.g. LY-CoV555) also target the RBM^28^. Despite strong affinity against WT-RBD (K_D_ = 5.8 nM), LY-CoV555 completely lost binding to NJ-RBD. Only 39% (7 out of 18) of the LY-CoV555 epitope residues were conserved between NJ-RBD and WT-RBD (**Extended Data Fig. 5b,g**). Class III antibodies (e.g. LY-CoV1404) target an epitope that is relatively conserved among VOCs/VOIs^29^. However, only 55% (11 of 20) of the LY-CoV1404 residues are conserved between NJ-RBD and WT-RBD. Consistently, NJ-RBD did not bind to LY-CoV1404, whereas WT-RBD showed strong binding (K_D_ = 4.4 nM) (**Extended Data Fig. 5c,h**). Class IV antibodies (e.g. CR3022) target a highly conserved cryptic epitope^30^. WT-RBD showed moderate binding to CR3022 (K_D_ = 40.8 nM), while NJ-RBD had a K_D_ weaker than 1000 nM. 83% of CR3022 epitope residues (25 of 30) are conserved between NJ-RBD and WT-RBD, which may explain the retained but very weak binding (**Extended Data Fig. 5d,i**). Class V antibodies (e.g. CC25.4) target an even more conserved epitope^31^,with only two substitutions between WT-RBD and NJ-RBD (K462Q and H519Q) (**Fig. 4b and Extended Data Fig. 5e**). Nevertheless, while CC25.4 still bound strongly to NJ-RBD (K_D_ = 4.4 nM), its affinity is 11-fold reduced compared to WT-RBD (K_D_ = 0.4 nM). Overall, the major known epitopes, even the conserved ones, are mutated in the NJ-RBD, resulting in substantial antibody evasion similar to that observed in the dominant Omicron subvariants.

**Fig. 4:**
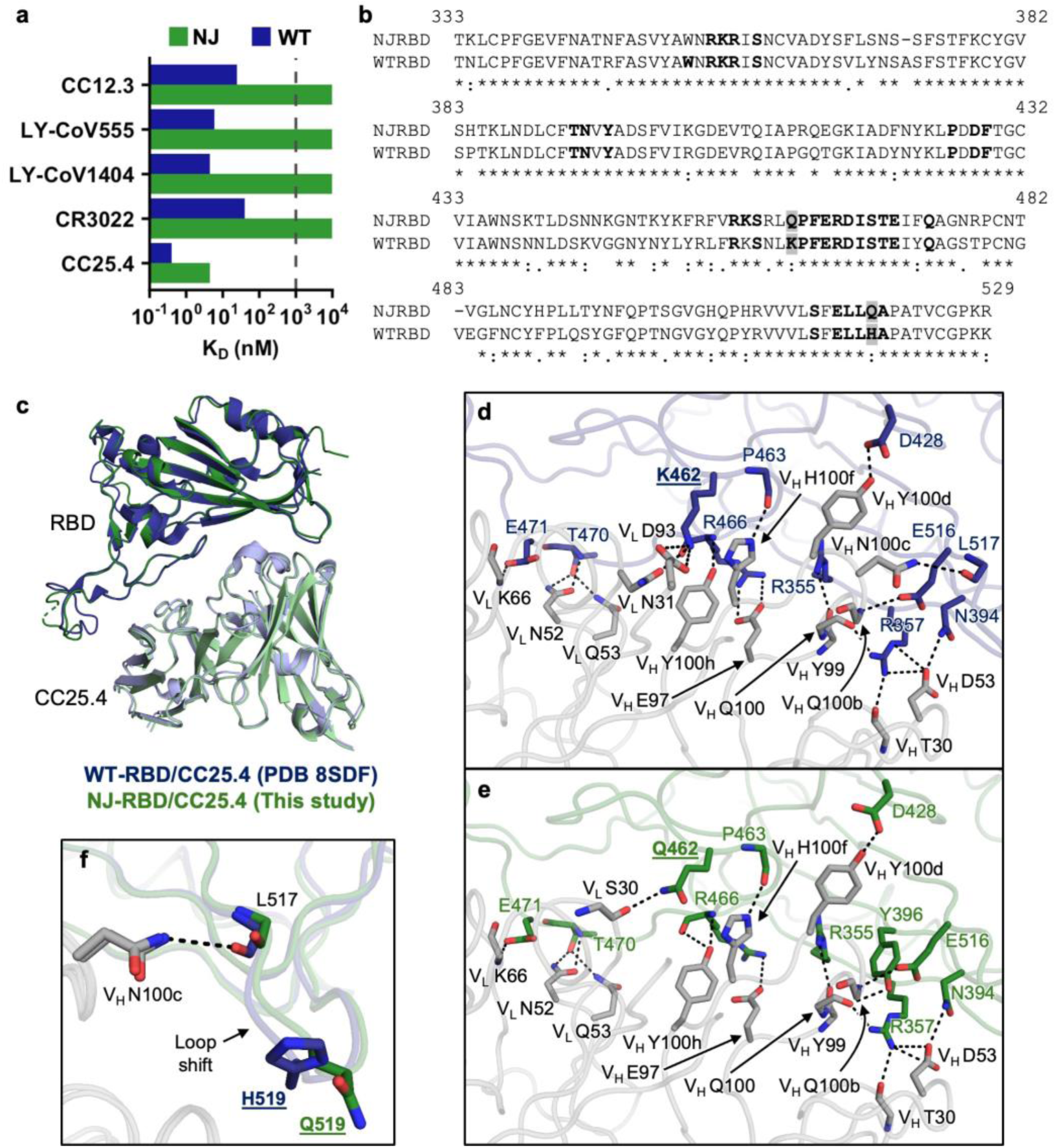
Structural comparison between NJ-RBD and WT-RBD in complex with CC25.4 Fab. **a**, Binding affinities of NJ-RBD (green) and WT-RBD (blue) with the indicated antibodies in Fab format by BLI. The dashed line indicates limit of detection. **b**, Sequence alignment of NJ-RBD and WT-RBD. CC25.4 epitope residues (BSA > 0 Å^2^ as calculated by PISA^54^) are in bold. Non-conserved epitope residues are highlighted in grey. **c**, NJ-RBD/CC25.4 (this study, green) with WT-RBD/CC25.4 (PDB 8SDF, blue)^31^ were aligned using their RBDs. Only antibody variable domains are shown for clarity. **d**,**e**, Molecular details of interactions (**d**) between WT-RBD and CC25.4, and between (**e**) NJ-RBD and CC25.4. RBD residues are numbered according to their positions on the SARS-CoV-2 WT spike protein sequence. Antibody residues have been numbered using the Kabat numbering scheme. H-bonds and salt bridges are represented by black dashed lines. Non-conserved epitope residues are shown in bold and underlined. **f**, H519Q substitution in the NJ-RBD introduced the loop shift and loss of the H-bond between CC25.4-V_H_ N100c and the main chain carbonyl of RBD-L517.

To investigate the detailed interactions between NJ-RBD and CC25.4, we determined a crystal structure of the NJ-RBD in complex with CC25.4 Fab at 2.8 Å resolution (**Fig. 4c and Extended Data Table 1**). Structural comparison with WT-RBD/CC25.4 complex (PDB 8SDF) revealed that the binding angles and positions of the antibody are nearly identical (Cα RMSD = 0.5 Å). The interface area and epitope residues are also very similar (**Fig. 4b,c**). A previous study found that CC25.4 interacts with the WT-RBD primarily through hydrophilic interactions^31^. Structural analysis showed that NJ-RBD also formed extensive and conserved hydrophilic interactions with CC25.4: RBD-R355, N394, D428, P463, E516, and E471 H-bonded with V_H_ Y99, D53, Y100d, H100f, Q100b, and V_L_ K66, respectively; RBD-R357 formed a salt bridge with V_H_ D53 and H-bonded with the V_H_ T30 main-chain and Q100; RBD-R466 formed a salt bridge with V_H_ E97 and H-bonded with V_H_ Y100h; RBD-T470 H-bonded with V_L_ N52 and Q53 (**Fig. 4d,e**). In addition to these conserved hydrophilic interactions, we observed that WT-RBD forms more H-bonds and salt bridges compared to NJ-RBD due to K462Q and H519Q in NJ-RBD (**Fig. 4d,e**). The K462Q substitution disrupted the salt bridge with V_L_ D93 and the H-bond with V_L_ N31, whereas the H519Q substitution caused a loop shift, resulting in the loss of the H-bond between L517 and V_H_ N100c (**Fig. 4f**). These differences likely explain why WT-RBD exhibited an 11-fold higher CC25.4-binding affinity than NJ-RBD (**Fig. 4a and Extended Data Fig. 5e**).

### The evolutionary landscape of NJ-RBD is shaped by epistasis

To understand the evolutionary landscape of NJ-RBD, we performed deep mutational scanning of its RBM (positions 437-507) using yeast display. Briefly, a site-saturation mutagenesis library of the NJ-RBM was constructed and displayed on the yeast surface. The impact of each amino acid substitution on hACE2 binding and expression levels was quantified using fluorescence-activated cell sorting (FACS) and next-generation sequencing (**see Methods, Fig. 5**). Pearson correlation coefficients of 0.98 (hACE2 binding) and 0.89 (expression) were obtained between two biological replicates, demonstrating the reproducibility of results (**Extended Data Fig. 6**). We found that the introduction of N-glycosylation sites by I468N and P499N increased RBD expression, consistent with previous findings that adding N-glycans to RBD help improve its expression^32^. However, P499N was highly detrimental to hACE2 binding (**Fig. 5a**), likely due to its proximity to the RBM.

**Fig. 5:**
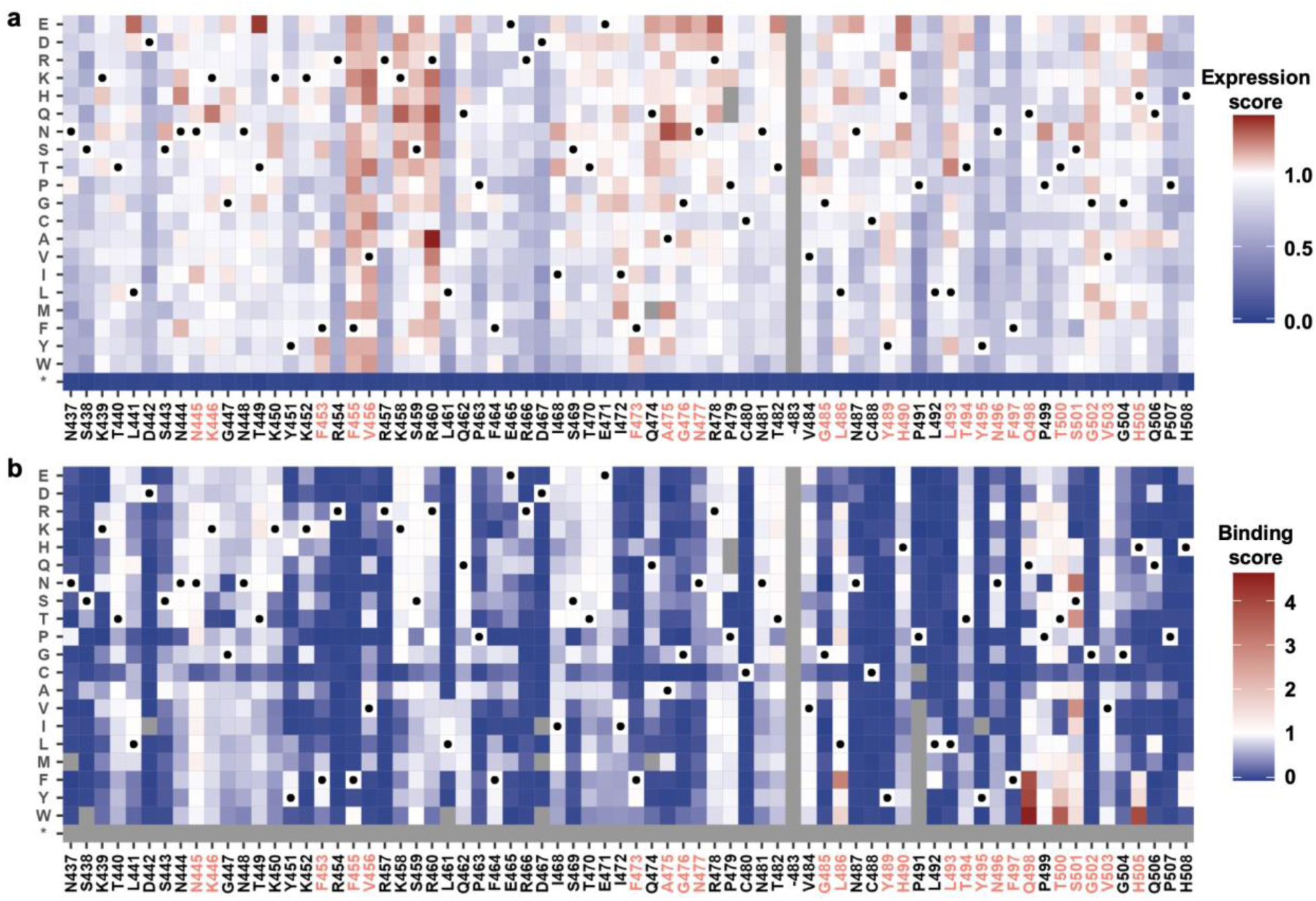
Deep mutational scanning of NJ-RBM. **a**,**b**, Heatmaps showing the effects of single substitutions on (**a**) RBM expression and (**b**) hACE2-binding affinity. The unmutated amino acids are indicated by a black circle. Substitutions with low occurrence frequency in mutant library are shown as grey in both panels a and b (**see Methods**). Substitutions with an expression score less than 0.5 are also shown as grey in panel b. * indicates substitution to a stop codon. The labels for residues participated in hACE2-binding are colored in red.

Despite already displaying exceptionally strong hACE2 affinity, the NJ variant could accommodate several additional substitutions that further enhance receptor binding (**Fig. 5b and Fig. 6a**). The hACE2 affinity-enhancing substitutions upon NJ-RBD were almost exclusively located within the 498-505 loop, which directly contacts hACE2 (**Fig 6a,c**). By contrast, most of these substitutions have negligible or even detrimental effects on hACE2 binding in dominant lineages^33–37^ (**Fig. 6a**). This differential effect on binding affinity may result from the substitution at residue 501. The RBD of NJ has S501, whereas N501 was present in that of variants earlier than Omicron and Y501 in that of Omicron and subvariants (**Fig. 6b,c**)^38^. RBD-N501 and RBD-Y501 interact with hACE2-Y41 via an H-bond and π-π stacking, respectively. They also H-bond with the main-chain of RBD-G496. None of these interactions were observed with RBD-S501 in NJ, which would increase the conformational flexibility of the 498-505 loop and hence the structural tolerance of substitutions in this region (**Fig. 6c**). Notably, the S501 substitution has been observed infrequently in circulating SARS-CoV-2, although reaching up to 4% occurrence in late 2023 (**Fig. 6d**). Overall, these observations illustrated how epistasis led to the unique evolutionary potential of NJ cryptic lineage.

**Fig. 6:**
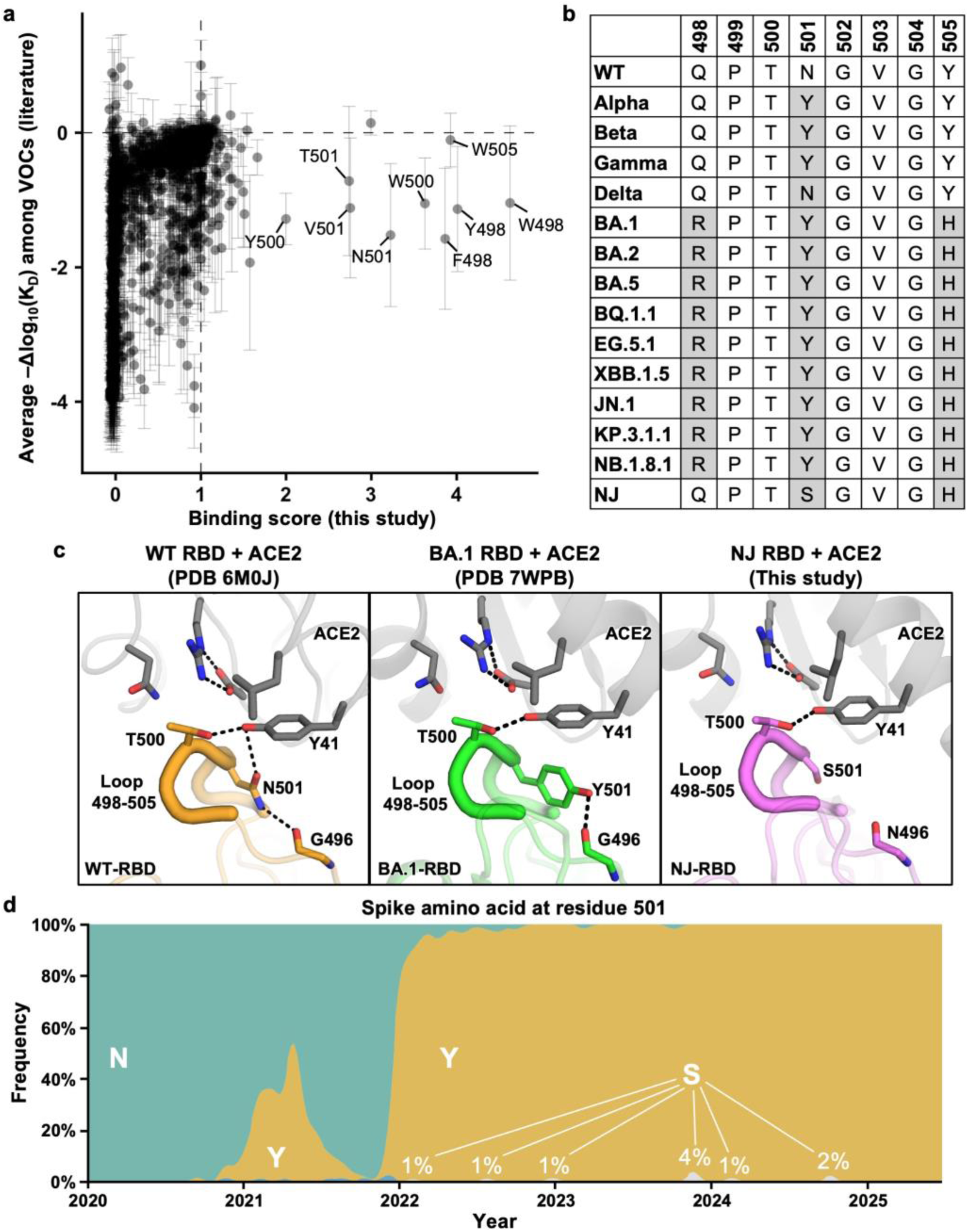
Epistasis in the 498-505 loop influences the evolutionary potential of NJ-RBD. **a**, Correlation between the hACE2-binding scores of individual mutations in NJ-RBD (this study) and their effects on hACE2-binding affinity in VOCs (mean ± SD) as determined previously by deep mutational scanning^33–37^. Dashed lines represented no effects on binding. **b**, Sequence alignment of the RBD residues 498-505 between the NJ cryptic lineage and VOCs. Amino acid variants differed from WT are highlighted in grey. **c**, Structures of hACE2 (grey) in complex with WT-RBD (PDB 6M0J, orange), BA.1-RBD (PDB 7WPB, green), and NJ-RBD (this study, pink)^18,38^. The 498-505 loop is shown as thick tube representation. H-bonds and salt bridges are represented by black dashed lines. **d**, Frequency of amino acid variants at RBD residue 501 of SARS-CoV-2 spike as of July 2025 was plotted using Nextstrain (https://nextstrain.org), which sources data from GISAID^55,56^.

## Discussion

During COVID-19 pandemic, wastewater viral sequencing has become as an effective surveillance tool and led to the discovery of cryptic SARS-CoV-2 lineages^6,7,9,13,39^, which raised many questions about SARS-CoV-2 biology. Previous studies have suggested that cryptic SARS-CoV-2 lineages may arise from immunocompromised individuals with persistent infections or from under-sampled reservoirs, including non-human hosts^6,7,9,13,39^. While the exact origin of the NJ cryptic lineage is currently unknown, there is a high chance that it emerged from a persistent human infection due to its enhanced binding affinity to hACE2 and extensive evasion against human antibodies. Additionally, our sequence analysis indicated that the NJ cryptic lineage likely replicated in the gastrointestinal tract. In fact, persistent infection by cryptic SARS-CoV-2 lineages seems to generally occur in the gastrointestinal tract^13^, potentially abolishing respiratory transmission and explaining why they are not widely circulating. However, the mechanisms of how cryptic SARS-CoV-2 lineages establishes persistent enteric infection have been largely elusive. Therefore, our study here provides important functional and structural insights into the biology of cryptic SARS-CoV-2 lineages.

Cryptic SARS-CoV-2 lineages usually carry numerous amino acid substitutions, which in turn can be categorized as either convergent or divergent^7,9^. Convergent evolution refers to the phenomenon where substitutions in cryptic lineages also independently emerged in circulating variants, primarily driven by shared selective pressures, such as receptor binding and antibody evasion. Specifically, NJ-RBD exhibits four convergent substitutions with Omicron or its subvariants, namely R403K, S477N, V483del, and Y505H. On the contrary, divergent evolution refers to the accumulation of unique substitutions that differ from those of circulating variants. The NJ cryptic lineage follows a distinct evolutionary path through unique substitutions, notably Q493L and G496N, which contribute to markedly enhanced hACE2-binding affinity, surpassing that of all tested Omicron subvariants. Nevertheless, substitutions that are currently unique to the NJ cryptic lineage may appear in future circulating variants, especially those involved in enhancing hACE2-binding affinity and antibody evasion. Therefore, the functional impact of each unique substitution in cryptic SARS-CoV-2 lineages, both on the spike and other part of the genome, warrants investigation to improve forecasting of SARS-CoV-2 evolution.

Persistent infection of cryptic SARS-CoV-2 lineages likely leads to intra-host co-evolution with the antibody repertoire, analogous to HIV infections^40,41^. Consistently, NJ-RBD showed extensive antibody evasion to different epitopes. Characterizing the antibody responses of individuals infected by cryptic SARS-CoV-2 lineages would benefit our understanding of how SARS-CoV-2 antibodies evolve to overcome escape mutations, which have important implications for developing broadly protective SARS-CoV-2 vaccines. However, such studies have been impossible due to the difficulties in identifying individuals persistently infected by cryptic SARS-CoV-2 lineages, even though narrowing down the origin of a given cryptic SARS-CoV-2 lineage to a particular toilet has proven feasible^6^. As more cryptic SARS-CoV-2 lineages continue to be discovered, future efforts should also focus on identifying and characterizing the corresponding infected individuals.

## Methods

### NCBI SRA screening and composite sequence generation

Sequences from NCBI SRA were processed and screened as previously described^13^. Briefly, NCBI SRA was searched with the term “SARS-CoV-2 wastewater”. Raw reads were downloaded and mapped to the SARS-CoV-2 genome (NC_045512) using Minimap2^42^ and the resulting sam file screened for the sequence “TACGATCGAGGGTACAGTG” in the mapped reads. The RBD region used in the analysis was assembled manually based on the sequences present in the samples from November 20 and 22, 2023.

For generating the composite sequence used for phylogenic analysis, we were able to include additional detections at later time points. We compared the sequences from sewer sheds containing cryptic lineage (SRR27717563, SRR27464193, SRR27023730, SRR27023734) to sequences from sewer sheds in the same state that did not contain the cryptic lineage (SRR27746262, SRR27563993, SRR27533535, SRR27391716, SRR27340850, SRR27240495, SRR27178282, SRR27050435, SRR26963081, SRR27717577, SRR27717596, SRR27464188, SRR27464300, SRR27464243, SRR27464240, SRR27318293, SRR27318288, SRR27228413, SRR27228429, SRR27152965, SRR27152979, SRR27023780) to determine polymorphisms present in NJ. The SRRs were processed using SAM Refiner, and the unique_seq and covar outputs processed by a custom script to determine mutations associated with each cryptic lineage by the following parameters: 1) the average sum abundance for the mutation must be 50× greater in the cryptic sewer sheds than in the non-cryptic sewer sheds; 2) a sum abundance of >10% of the maximum sum abundance of the most abundant polymorphism for a cryptic-specific mutation from those sewer shed samples. To account for mutations prevalent in both circulating and cryptic lineages, any polymorphism appearing at least 75% of the time in the same sequence read as a cryptic-specific polymorphism was considered to be “linked” and present in that lineage. To create a composite genome sequence, the NC_045512 sequence was changed to have the nucleotide sequences from the above polymorphisms, and any position that did not have a coverage depth of at least five from the reads containing the cryptic-specific mutations was changed to “N”. To generate the phylogenic tree, the composite genome was run through the Nextclade web interface (https://clades.nextstrain.org)^43^.

### Expression and purification of human ACE2

For X-ray crystallography, human ACE2 (residues 19-615, GenBank: BAJ21180.1) was flanked with an N-terminal signal peptide and a C-terminal His_6_ tag and codon optimized in phCMV3 expression vector. Expi293S cells (Thermo Fisher Scientific) were then transfected using the ExpiFectamine 293 Transfection Kit (Thermo Fisher Scientific) according to the manufacturer’s instructions. Supernatants were harvested and proteins were purified by Ni Sepharose excel resin (Cytiva), eluted with 300 mM imidazole. For SPR binding, human ACE2 constructs were fused with a human IgG1 Fc domain. Expi293F cells (Thermo Fisher Scientific) were transfected using the ExpiFectamine 293 Transfection Kit (Thermo Fisher Scientific) according to the manufacturer’s instructions. Supernatants were harvested and ACE2-Fc proteins were purified with a HiTrap Protein A HP column (Cytiva) and eluted with 50 mM sodium acetate, pH 3.5. Protein samples were further purified with size exclusion (SEC) and buffer exchanged into 20 mM Tris-HCl, 150 mM NaCl, pH 7.4.

### Expression and purification of Fab proteins

The heavy and light chains of the Fabs (CC12.3, LY-CoV555, LY-CoV1404, CR3022, and CC25.4) were cloned into the phCMV3 vector. The plasmids were transiently co-transfected into ExpiCHO cells at a ratio of 2:1 (heavy chain to light chain) using ExpiFectamine CHO Reagent (Thermo Fisher Scientific) according to the manufacturer’s instructions. The supernatant was collected at 7 days post-transfection. The Fab was purified with a CaptureSelect CH1-XL Pre-packed Column (Thermo Fisher Scientific), followed by SEC and buffer exchanged 20 mM Tris-HCl, 150 mM NaCl, pH 7.4.

### Expression and purification of RBDs

The receptor-binding domains (RBDs) (residues 333-528) of the SARS-CoV-2 spike (S) protein of all variants (wild-type, Omicron BA.1, BA.4, BQ.1.1, XBB.1.5, XBB.1.16, JN.1, KP.2, KP.3) and cryptic lineage NJ were cloned into a customized pFastBac vector. The RBD constructs were fused with an N-terminal gp67 signal peptide and a C-terminal His_6_ tag. Recombinant bacmid DNA was generated using the Bac-to-Bac system (Thermo Fisher Scientific). Baculovirus was generated by transfecting purified bacmid DNA into Sf9 cells using FuGENE HD (Promega), and subsequently used to infect suspension cultures of Sf9 cells at an MOI of 5 to 10. Infected Sf9 cells were incubated at 28°C with shaking at 110 r.p.m. for 72 h for protein expression. The supernatant was then concentrated using a 10 kDa MW cutoff Centramate cassette (Pall Corporation). The RBD proteins were purified by Ni-NTA, followed by size exclusion chromatography, and buffer exchanged into 20 mM Tris-HCl, 150 mM NaCl, pH 7.4.

### Surface plasmon resonance (SPR) binding assays

SPR measurements were conducted on Biacore S200 instrument (Cytiva). TBST (TBS with 0.005% Tween-20) buffer was used as running buffer and to reconstitute all proteins. Fc-tagged hACE2 were captured on the Sensor Chip Protein A (Cytiva). The capture level was targeted to 2,500 response units. A blank flow cell was treated in parallel without hACE2 injection. After immobilization, multicycle kinetic measurements were conducted on each recombinant RBD analyte. Analytes at 300 nM, 100 nM, 33.3 nM, 11.1 nM, 3.7 nM, and 0 nM were injected at 30 μL/min for 120 s and followed by 240 s dissociation. After each cycle, 2 steps of 30 s regeneration with 10 mM glycine buffer, pH 1.7, were used to achieve a flat baseline signal. Final SPR sensorgrams were collected by reference channel subtraction and zero concentration subtraction. The resulting data were fitted to a 1:1 binding model using Biacore evaluation software.

### Biolayer interferometry (BLI) binding assays

Binding assays were performed by biolayer interferometry (BLI) using an Octet Red instrument (FortéBio). His_6_-tagged RBDs at 20 μg/mL in 1× kinetics buffer (1× PBS, pH 7.4, 0.01% BSA, and 0.002% Tween 20) were loaded onto NI-NTA sensors, and then incubated with 33.3 nM, 100 nM, and 300 nM of Fabs. The assay consisted of five steps: 1) baseline: 60 s with 1× kinetics buffer; 2) loading: 90 s with His_6_-tagged RBDs; 3) baseline: 90 s with 1× kinetics buffer; 4) association: 180 s with Fabs; and 5) dissociation: 180 s with 1× kinetics buffer. For estimating the K_D_ values, a 1:1 binding model was used.

### Crystal structure determination

For crystallization, purified NJ-RBD and hACE2, as well as NJ-RBD and CC25.4 Fab, were mixed at an equimolar ratio and incubated overnight at 4°C. A VSRRLP variant of the elbow region was used to reduce the conformational flexibility between the heavy chain constant and variable domains of Fab CC25.4^44^. NJ-RBD/hACE2 complexes (12 mg/mL) and NJ-RBD/CC25.4 Fab complexes (12.5 mg/mL) were screened for crystallization with the 384 conditions of the JCSG Core Suite (QIAGEN) on our custom-designed robotic CrystalMation system (Rigaku) at Scripps Research by the vapor diffusion method in sitting drops containing 0.1 μL of protein and 0.1 μL of reservoir solution. Diffraction-quality crystals were obtained in the following conditions:

NJ-RBD/hACE-2: 0.1 M Tris, pH 7, 0.2 M magnesium chloride, and 10% (w/v) polyethylene glycol 8000 at 20°C.
NJ-RBD/CC25.4 Fab: 0.1 M sodium acetate, pH 4.6, and 2 M ammonium sulfate at 20°C.

Crystals appeared on day 14 and were harvested on day 25 by soaking in reservoir solution supplemented with 20% (v/v) ethylene glycol as cryoprotectant. The crystals were then flash-cooled and stored in liquid nitrogen until data collection. Diffraction data were collected at cryogenic temperature (100 K) at Stanford Synchrotron Radiation Lightsource (SSRL) beamline 12-1 with a beam wavelength of 0.97946 Å. Diffraction data were processed with HKL2000^45^. Structures were solved by molecular replacement using PHASER^46^ and two structures (PDB 6M0J and 8SDF) as initial model^18,31^. Iterative model building and refinement were carried out in COOT^47^ and PHENIX^48^, respectively.

### Mutant library construction

NJ-RBM yeast display plasmid, pCTcon2_NJ_RBM, was constructed by cloning the coding sequences of (from N-terminal to C-terminal, all in-frame) Aga2 secretion signal, NJ-RBD, HA tag, and Aga2p, into the pCTcon2 vector^49^. To generate the linearized vector for the library, pCTcon2_NJ_RBM was used as a template for PCR using 5’-TTG AAG GTA CTG AAG GAT GAA TTA CTC-3’ and 5’-AGG AAC TGA CAA CTA TAT GCG AGC AAA-3’ as primers. The PCR product was then gel-purified. For insert generation, two batches of PCRs were performed, followed by overlapping PCRs. The first batch of PCRs consisted of 9 reactions, each with an equal molar mix of eight forward primers, as the forward primer and a universal reverse primer 5’-CGA TTC TAA AGT TGG TGA GGG GAT TTG CTC GCA TAT AGT TGT CAG TTC CT-3’. The forward primers for the first batch of PCRs are listed in **Extended Data Table 2**. These forward primers were named as cassetteX_N, in which X represents the cassette ID and N represents the primer number. Forward primers with the same cassette ID were mixed at equal molar ratios and used in the same single PCR. The second batch of PCRs consisted of another 9 reactions, each with a universal forward primer 5’-TGT GTG GCA GAT TAC TCG TTC CTG AGT AAT TCA TCC TTC AGT ACC TTC AA-3’ and a unique reverse primer as listed in **Extended Data Table 3**. Subsequently, nine overlapping PCRs were performed using the universal forward and reverse primers. For each overlapping PCR, the template was a mixture of 10 ng each of the corresponding products from the first and second batches of PCRs. The complete insert was an equal molar mix of the products of these nine overlapping PCRs. All PCRs were performed using PrimeSTAR Max polymerase (Takara Bio) according to the manufacturer’s instructions. PCR products were purified using Monarch DNA Gel Extraction Kit (New England Biolabs).

### Yeast transformation

Yeast cells were transformed by electroporation following a previously described protocol^50^. Briefly, *Saccharomyces cerevisiae* EBY100 cells (American Type Culture Collection) were grown in YPD medium (1% w/v yeast nitrogen base, 2% w/v peptone, 2% w/v D(+)-glucose) overnight at 30°C with shaking at 225 rpm until OD_600_ reached 3. Then, an aliquot of overnight culture was grown in 100 mL YPD media, with an initial OD_600_ of 0.3, shaking at 225 rpm at 30 °C. Once OD_600_ reached 1.6, yeast cells were collected by centrifugation at 1700 × *g* for 3 min at room temperature. Media were removed and the cell pellet was washed twice with 50 mL ice-cold water, and then once with 50 mL of ice-cold electroporation buffer (1 M sorbitol, 1 mM calcium chloride). Cells were resuspended in 20 mL conditioning media (0.1 M lithium acetate, 10 mM dithiothreitol) and shaken at 225 rpm at 30°C. Cells were collected via centrifugation at 1700 × *g* for 3 min at room temperature, washed once with 50 mL ice-cold electroporation buffer, resuspended in electroporation buffer to reach a final volume of 1 mL, and kept on ice. 5 µg of the complete insert and 4 µg of the purified linearized vector were added into 400 µL of conditioned yeast. The mixture was transferred to a pre-chilled BioRad GenePulser cuvette with 2 mm electrode gap and kept on ice for 5 min until electroporation. Cells were electroporated at 2.5 kV and 25 µF, achieving a time constant between 3.7 and 4.1 ms. Electroporated cells were transferred into 4 mL of YPD media supplemented with 4 mL of 1 M sorbitol and incubated at 30°C with shaking at 225 rpm for 1 h. Cells were collected via centrifugation at 1700 × *g* for 3 min at room temperature, resuspended in 0.6 mL SD-CAA medium (2% w/v D-glucose, 0.67% w/v yeast nitrogen base with ammonium sulfate, 0.5% w/v casamino acids, 0.54% w/v Na_2_HPO_4_, 0.86% w/v NaH_2_PO_4_·H_2_O, all dissolved in deionized water), plated onto SD-CAA plates (2% w/v D-glucose, 0.67% w/v yeast nitrogen base with ammonium sulfate, 0.5% w/v casamino acids, 0.54% w/v Na_2_HPO_4_, 0.86% w/v NaH_2_PO_4_·H_2_O, 18.2% w/v sorbitol, 1.5% w/v agar, all dissolved in deionized water) and incubated at 30 °C for 40 h. Colonies were then collected in SD-CAA medium, centrifuged at 1700 × *g* for 5 min at room temperature, and resuspended in SD-CAA medium with 15% v/v glycerol such that OD_600_ was 50. Glycerol stocks were stored at −80°C until use.

### Fluorescence-activated cell sorting (FACS) of yeast display mutant library

100 µL glycerol stock of the yeast display library was recovered in 50 mL SD-CAA medium by incubating at 27 °C with shaking at 250 rpm until OD_600_ reached between 1.5 and 2.0. Then 15 mL of the yeast culture was harvested and pelleted via centrifugation at 4000 × *g* at 4°C for 5 min. The supernatant was discarded, and SGR-CAA (2% w/v galactose, 2% w/v raffinose, 0.1% w/v D-glucose, 0.67% w/v yeast nitrogen base with ammonium sulfate, 0.5% w/v casamino acids, 0.54% w/v Na_2_HPO_4_, 0.86% w/v NaH_2_PO_4_·H_2_O, all dissolved in deionized water) was added to make up the volume to 50 mL. The yeast culture was then transferred to a baffled flask and incubated at 18°C with shaking at 250 rpm. Once OD_600_ reached between 1.3 and 1.6, 1 mL of yeast culture was harvested and pelleted via centrifugation at 4000 × *g* at 4°C for 5 min. The pellet was subsequently washed with 1 mL of 1× PBS twice. After the final wash, cells were resuspended in 1 mL of 1× PBS.

For expression sort, APC anti-HA.11 (epitope 16B12, BioLegend, Cat. No. 901524) that was buffer-exchanged into 1× PBS was added to the cells at a final concentration of 1 µg/mL. For binding sort, Fc-tagged human ACE2 was added to washed cells at a final concentration of 1 µM. A negative control was set up with nothing added to the PBS-resuspended cells. Samples were incubated overnight at 4°C with rotation. The yeast pellet was then washed twice in 1× PBS. After the last wash, cells were resuspended in 1 ml of 1× PBS. For the binding sort, PE anti-human IgG Fc antibody (clone M1310G05, BioLegend, Cat. No. 410708) that was buffer-exchanged into 1× PBS was added to yeast at a final concentration of 1 µg/mL. Cells were incubated at 4°C for 1 h with rotation. Then, the yeast pellet was washed twice in 1× PBS and resuspended in FACS tubes containing 2 mL 1× PBS. Using a BD FACSMelody cell sorter (BD Biosciences) and FACS Diva software v9.0 (BD Biosciences), single yeast cells were gated by forward scatter (FSC) and side scatter (SSC). Single cells were further partitioned into four bins: bin 1 captured 99% of unstained cells, and bins 2-4 split the remaining library fraction into tertiles. Singleton APC-positive cells and PE-positive cells were collected in 1 mL of SD-CAA containing 1× penicillin/streptomycin for expression and binding sort, respectively. The post-sort samples were subsequently centrifuged at 4000 × *g* at 20°C for 30 min. The supernatant was discarded. Subsequently, the pellet was resuspended in 3 mL of SD-CAA and were grown overnight at 30°C. Each replicate was prepared and sorted independently. Plasmids were purified from the unsorted yeast display library (input) as well as two replicates of post-sort yeast samples using Zymo Yeast Miniprep kits (Zymo Research) following the manufacturer’s protocol. The extracted plasmids were stored at −20°C until used.

### Next-generation sequencing of the NJ-RBM mutant library

The NJ-RBM library was subsequently amplified by PCR using recovery forward primer 5’-CAC TCT TTC CCT ACA CGA CGC TCT TCC GAT CTG ACT TTA CCG GTT GTG TCA TTG CA-3’ and recovery reverse primer 5’-GAC TGG AGT TCA GAC GTG TGC TCT TCC GAT CTG TTC GAA TGA GAG AAC CAC CAC-3’. Subsequently, adapters containing sequencing barcodes were appended to the amplicon using primers 5′-AAT GAT ACG GCG ACC ACC GAG ATC TAC ACX XXX XXX XAC ACT CTT TCC CTA CAC GAC GCT-3′, and 5′-CAA GCA GAA GAC GGC ATA CGA GAT XXX XXX XXG TGA CTG GAG TTC AGA CGT GTG CT-3′. Positions annotated by an “X” represented the nucleotides for the index sequence. All PCRs were performed using Q5 High-Fidelity DNA polymerase (New England Biolabs) according to the manufacturer’s instructions. PCR products were purified using Monarch DNA Gel Extraction Kit (New England Biolabs). The final PCR products of each replicate was prepared individually and submitted for next generation sequencing using MiSeq Bulk v2 PE250 (Illumina).

### Analysis of sequencing data for deep mutational scanning

The sequencing data was initially obtained in FASTQ format and subsequently analyzed using a customized python Snakemake pipeline^51^. Briefly, a reference mutational library containing all possible amino acid substitutions in the gene of interest was generated using Biopython^52^ and library primers listed in **Table S2**. The sequencing reads were aligned to this reference library using BWA 0.8.17^53^. Reads were retained only if both forward and reverse sequences perfectly matched the reference; otherwise, they were discarded. The number of reads corresponding to each variant in each sample was counted. A pseudocount of 1 was added to the final count to avoid division by zero in downstream analysis. The fluorescence intensity of each variant *var* was used as a proxy for the binding and expression enrichment values was computed as follows:

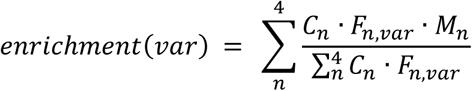

where *C*_*n*_ is the number of cells sorted in each gate, *F*_*n*,*var*_ is the frequency of each variant in each gate, and *M*_*n*_ is the mean fluorescence intensity (MFI) for each gate. The binding and expression scores for each variant were further computed from the enrichment values as follows:

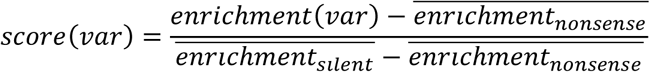

where 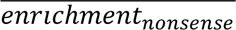 is the average enrichment value of the of the nonsense mutations and 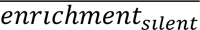 is the average enrichment value of the silent mutations. The final score for each variant *var* is the average of two biological replicates. Variants with a read count less than 10 in the input were discarded for downstream analysis.

## Data availability

Raw sequencing data have been submitted to the NIH Short Read Archive under accession numbers: BioProject PRJNA1237868. The X-ray coordinates and structure factors have been deposited to the RCSB Protein Data Bank under accession codes: 9Z1F and 9Z1G.

## Code availability

Custom python scripts for all analyses have been deposited to: https://github.com/nicwulab/NJ-RBM_DMS.

## Acknowledgements

The Wilson lab is grateful to the staff of the Stanford Synchrotron Radiation Lightsource (SSRL) beamlines 12-1 for assistance. Use of the Stanford Synchrotron Radiation Lightsource, SLAC National Accelerator Laboratory, is supported by the U.S. Department of Energy, Office of Science, Office of Basic Energy Sciences under Contract No. DE-AC02-76SF00515. The SSRL Structural Molecular Biology Program is supported by the DOE Office of Biological and Environmental Research, and by the National Institutes of Health, National Institute of General Medical Sciences (P30GM133894). We thank Henry Tien for technical support with the crystallization robot, Robyn Stanfield for assistance in data collection, Marc-André Elsliger with computation, Jeanne Matteson and Beverly Ellis for their contribution to mammalian cell culture. Wea also thank the Roy J. Carver Biotechnology Center at the University of Illinois at Urbana-Champaign for assistance with fluorescence-activated cell sorting and next-generation sequencing. This study was supported by Gates Foundation INV-004923 (I.A.W.), the Vallee Scholars Program (N.C.W.), and the National Institute of Allergy and Infectious Diseases R01 AI190286 (I.A.W. and M.Y.), R01 AI165475 (N.C.W.), and DP2 AT011966 (N.C.W.).

## Author information Contributions

Z.F., M.C.J, I.A.W., M.Y., and N.C.W. conceived the study. I.A.W., M.Y., and N.C.W. supervised the study. Z.F. acquired and analyzed X-ray crystallographic data. Z.F. and D.Z. performed binding assays and interpreted data. Q.W.T. W.L., K.J.M., and E.K.S. performed the deep mutational scanning and data analysis. B.A.I., D.A.G., and M.C.J. performed wastewater sequencing and data analysis. Z.F., M.C.J., M.Y., and N.C.W. wrote the paper, and all authors reviewed and edited the paper. The funding was secured by M.C.J, M.Y., I.A.W., and N.C.W.

