## Supplementary material for "A highly divergent cryptic SARS-CoV-2 lineage exhibits strong receptor binding and immune evasion": Tables S1-S3 and Figures S1-S6

### Extended Data Table 1. X-ray data collection and refinement statistics

| **Data collection** | NJ-RBD + hACE2 | NJ-RBD + CC25.4 |
|---|---|---|
| Beamline | SSRL BL 12-1 | SSRL BL 12-1 |
| Wavelength (Å) | 0.97946 | 0.97946 |
| Space group | P 1 $2_1$ 1 | I 2 |
| Unit cell parameters | | |
| a, b, c (Å) | 81.6, 123.0, 110.4 | 126.4, 239.5, 152.2 |
| α, β, γ (°) | 90, 91.1, 90 | 90, 113.9, 90 |
| Resolution (Å) [a] | 50.0-2.50 (2.54-2.50) | 50.0-2.80 (2.85-2.80) |
| Unique reflections [a] | 73,735 (3,725) | 96,329 (4,834) |
| Redundancy [a] | 6.9 (6.6) | 6.9 (6.4) |
| Completeness (%) [a] | 98.1 (98.8) | 98.5 (99.2) |
| $<I/\sigma_I>$ [a] | 17.0 (1.1) | 14.5 (1.5) |
| $R_{sym}$[b] (%) [a] | 11.7 (>100) | 20.0 (>100) |
| $R_{pim}$[b] (%) [a] | 4.7 (86.1) | 8.1 (94.4) |
| $CC_{1/2}$[c] (%) [a] | 99.1 (49.7) | 99.0 (63.3) |
| **Refinement statistics** | | |
| Resolution (Å) | 36.7-2.50 | 38.7-2.80 |
| Reflections (work) | 61,736 | 85,600 |
| Reflections (test) | 1,992 | 2,000 |
| $R_{cryst}$[d] / $R_{free}$[e] (%) | 22.0/26.6 | 22.5/25.9 |
| Copies of complex per ASU | 2 | 3 |
| No. of atoms | 13,349 | 18,171 |
| Macromolecules | 13,232 | 17,979 |
| Ligands [f] | 32 | 123 |
| Waters | 85 | 69 |
| Average *B*-values (Å$^2$) | 44 | 57 |
| Macromolecules | 44 | 57 |
| Ligands [f] | 70 | 78 |
| Waters | 30 | 38 |
| Wilson *B*-value (Å$^2$) | 37 | 53 |
| **RMSD from ideal geometry** | | |
| Bond length (Å) | 0.002 | 0.002 |
| Bond angle (°) | 0.53 | 0.56 |
| **Ramachandran statistics (%)** [g] | | |
| Favored | 97.3 | 96.2 |
| Outliers | 0.1 | 0.1 |
| **PDB code** | 9Z1F | 9Z1G |

[a] Numbers in parentheses refer to the highest resolution shell.

[b] $R_{sym} = \Sigma_{hkl} \Sigma_i | I_{hkl,i} - <I_{hkl}> | / \Sigma_{hkl} \Sigma_i I_{hkl,i}$ and $R_{pim} = \Sigma_{hkl} (1/(n-1))^{1/2} \Sigma_i | I_{hkl,i} - <I_{hkl}> | / \Sigma_{hkl} \Sigma_i I_{hkl,i}$, where $I_{hkl,i}$ is the scaled intensity of the $i^{th}$ measurement of reflection h, k, l, $<I_{hkl}>$ is the average intensity for that reflection, and *n* is the redundancy.

[c] $CC_{1/2}$ = Pearson correlation coefficient between two random half datasets.

[d] $R_{cryst} = \Sigma_{hkl} | F_o - F_c | / \Sigma_{hkl} | F_o | \times 100$, where $F_o$ and $F_c$ are the observed and calculated structure factors, respectively.

[e] $R_{free}$ was calculated as for $R_{cryst}$, but on a test set comprising ~2.4%-5.1% of the data excluded from refinement.

[f] Bound ligands are ethylene glycol, zinc ion, and sulfate molecules.

[g] From MolProbity[1].

**Extended Data Table 2. Sequences of the forward primers for constructing NJ-RBM mutant library.**

| Primer ID | Sequence (5' to 3') |
|---|---|
| Cassette1_1 | ACCGGTTGTGTCATTGCATGGNNKTCTAAGACGCTTGACAGCAATAATAAGGGCAATACC |
| Cassette1_2 | ACCGGTTGTGTCATTGCATGGAATNNKAAAACCCTTGACAGCAATAATAAGGGCAATACC |
| Cassette1_3 | ACCGGTTGTGTCATTGCATGGAACTCTNNKACCTTAGACAGCAATAATAAGGGCAATACC |
| Cassette1_4 | ACCGGTTGTGTCATTGCATGGAATAGTAAGNNKTTAGACAGCAATAATAAGGGCAATACC |
| Cassette1_5 | ACCGGTTGTGTCATTGCATGGAACTCTAAAACGNNKGATAGCAATAATAAGGGCAATACC |
| Cassette1_6 | ACCGGTTGTGTCATTGCATGGAACAGTAAGACCCTTNNKAGCAATAATAAGGGCAATACC |
| Cassette1_7 | ACCGGTTGTGTCATTGCATGGAATTCTAAAACGTTAGACNNKAATAATAAGGGCAATACC |
| Cassette1_8 | ACCGGTTGTGTCATTGCATGGAACAGTAAGACGTTAGATAGCNNKAATAAGGGCAATACC |
| Cassette2_1 | AGTAAAACGCTTGACAGCAATNNKAAAGGTAATACCAAGTACAAGTTCCGTTTCGTGAGG |
| Cassette2_2 | AGTAAAACGCTTGACAGCAATAACNNKGGCAACACCAAGTACAAGTTCCGTTTCGTGAGG |
| Cassette2_3 | AGTAAAACGCTTGACAGCAATAACAAGNNKAATACAAAGTACAAGTTCCGTTTCGTGAGG |
| Cassette2_4 | AGTAAAACGCTTGACAGCAATAATAAAGGCNNKACAAAGTACAAGTTCCGTTTCGTGAGG |
| Cassette2_5 | AGTAAAACGCTTGACAGCAATAATAAGGGTAACNNKAAGTACAAGTTCCGTTTCGTGAGG |
| Cassette2_6 | AGTAAAACGCTTGACAGCAATAACAAAGGTAACACANNKTACAAGTTCCGTTTCGTGAGG |
| Cassette2_7 | AGTAAAACGCTTGACAGCAATAACAAGGGTAACACCAAANNKAAGTTCCGTTTCGTGAGG |
| Cassette2_8 | AGTAAAACGCTTGACAGCAATAACAAGGGCAACACAAAATACNNKTTCCGTTTCGTGAGG |
| Cassette3_1 | AAGGGCAATACCAAGTACAAGNNKCGATTTGTGAGGAAGAGCCGCTTGCAGCCCTTTGAA |
| Cassette3_2 | AAGGGCAATACCAAGTACAAGTTTNNKTTCGTAAGGAAGAGCCGCTTGCAGCCCTTTGAA |
| Cassette3_3 | AAGGGCAATACCAAGTACAAGTTTCGTNNKGTGCGAAAGAGCCGCTTGCAGCCCTTTGAA |
| Cassette3_4 | AAGGGCAATACCAAGTACAAGTTCCGATTCNNKCGAAAGAGCCGCTTGCAGCCCTTTGAA |
| Cassette3_5 | AAGGGCAATACCAAGTACAAGTTCCGTTTTGTANNKAAGAGCCGCTTGCAGCCCTTTGAA |
| Cassette3_6 | AAGGGCAATACCAAGTACAAGTTTCGATTTGTACGANNKAGCCGCTTGCAGCCCTTTGAA |
| Cassette3_7 | AAGGGCAATACCAAGTACAAGTTTCGATTCGTGCGAAAANNKCGCTTGCAGCCCTTTGAA |
| Cassette3_8 | AAGGGCAATACCAAGTACAAGTTTCGTTTTGTAAGGAAAAGCNNKTTGCAGCCCTTTGAA |
| Cassette4_1 | CGTTTCGTGAGGAAGAGCCGCNNKCAACCTTTTGAAAGGGACATCTCAACTGAAATTTTT |
| Cassette4_2 | CGTTTCGTGAGGAAGAGCCGCTTANNKCCCTTCGAAAGGGACATCTCAACTGAAATTTTT |
| Cassette4_3 | CGTTTCGTGAGGAAGAGCCGCTTACAGNNKTTTGAGAGGGACATCTCAACTGAAATTTTT |
| Cassette4_4 | CGTTTCGTGAGGAAGAGCCGCTTGCAACCCNNKGAGAGGGACATCTCAACTGAAATTTTT |
| Cassette4_5 | CGTTTCGTGAGGAAGAGCCGCTTGCAGCCTTTCNNKAGGGACATCTCAACTGAAATTTTT |
| Cassette4_6 | CGTTTCGTGAGGAAGAGCCGCTTACAACCTTTCGAGNNKGACATCTCAACTGAAATTTTT |
| Cassette4_7 | CGTTTCGTGAGGAAGAGCCGCTTACAACCCTTTGAGCGANNKATCTCAACTGAAATTTTT |
| Cassette4_8 | CGTTTCGTGAGGAAGAGCCGCTTACAGCCTTTTGAACGAGACNNKTCAACTGAAATTTTT |
| Cassette5_1 | CAGCCCTTTGAAAGGGACATCNNKACCGAGATTTTTCAAGCGGGTAATCGCCCATGCAAT |
| Cassette5_2 | CAGCCCTTTGAAAGGGACATCTCTNNKGAAATCTTTCAAGCGGGTAATCGCCCATGCAAT |
| Cassette5_3 | CAGCCCTTTGAAAGGGACATCTCTACTNNKATTTTCCAAGCGGGTAATCGCCCATGCAAT |
| Cassette5_4 | CAGCCCTTTGAAAGGGACATCTCAACCGAANNKTTCCAAGCGGGTAATCGCCCATGCAAT |
| Cassette5_5 | CAGCCCTTTGAAAGGGACATCTCAACTGAGATCNNKCAAGCGGGTAATCGCCCATGCAAT |
| Cassette5_6 | CAGCCCTTTGAAAGGGACATCTCTACCGAGATCTTCNNKGCGGGTAATCGCCCATGCAAT |
| Cassette5_7 | CAGCCCTTTGAAAGGGACATCTCTACCGAAATTTTTCAGNNKGGTAATCGCCCATGCAAT |
| Cassette5_8 | CAGCCCTTTGAAAGGGACATCTCTACTGAGATCTTTCAGGCGNNKAATCGCCCATGCAAT |
| Cassette6_1 | ACTGAAATTTTTCAAGCGGGTNNKCGACCTTGCAATACCGTGGGCCTTAATTGCTACCAC |
| Cassette6_2 | ACTGAAATTTTTCAAGCGGGTAACNNKCCATGTAATACCGTGGGCCTTAATTGCTACCAC |
| Cassette6_3 | ACTGAAATTTTTCAAGCGGGTAACCGCNNKTGCAACACCGTGGGCCTTAATTGCTACCAC |
| Cassette6_4 | ACTGAAATTTTTCAAGCGGGTAATCGACCANNKAACACCGTGGGCCTTAATTGCTACCAC |
| Cassette6_5 | ACTGAAATTTTTCAAGCGGGTAATCGCCCTTGTNNKACCGTGGGCCTTAATTGCTACCAC |
| Cassette6_6 | ACTGAAATTTTTCAAGCGGGTAACCGACCTTGTAACNNKGTGGGCCTTAATTGCTACCAC |
| Cassette6_7 | ACTGAAATTTTTCAAGCGGGTAACCGACCATGCAACACANNKGGCCTTAATTGCTACCAC |
| Cassette6_8 | ACTGAAATTTTTCAAGCGGGTAACCGCCCTTGTAATACAGTGNNKCTTAATTGCTACCAC |
| Cassette7_1 | CGCCCATGCAATACCGTGGGCNNKAACTGTTACCACCCCTTGCTGACATACAACTTTCAA |
| Cassette7_2 | CGCCCATGCAATACCGTGGGCTTANNKTGCTATCACCCCTTGCTGACATACAACTTTCAA |
| Cassette7_3 | CGCCCATGCAATACCGTGGGCTTAAATNNKTACCATCCCTTGCTGACATACAACTTTCAA |
| Cassette7_4 | CGCCCATGCAATACCGTGGGCCTTAACTGCNNKCATCCCTTGCTGACATACAACTTTCAA |
| Cassette7_5 | CGCCCATGCAATACCGTGGGCCTTAATTGTTATNNKCCCTTGCTGACATACAACTTTCAA |
| Cassette7_6 | CGCCCATGCAATACCGTGGGCTTAAACTGTTATCATNNKTTGCTGACATACAACTTTCAA |
| Cassette7_7 | CGCCCATGCAATACCGTGGGCTTAAACTGCTACCATCCTNNKCTGACATACAACTTTCAA |
| Cassette7_8 | CGCCCATGCAATACCGTGGGCTTAAATTGTTATCACCCTTTGNNKACATACAACTTTCAA |
| Cassette8_1 | AATTGCTACCACCCCTTGCTGNNKTATAATTTTCAACCCACTAGTGGTGTCGGTCATCAA |
| Cassette8_2 | AATTGCTACCACCCCTTGCTGACCNNKAACTTCCAACCCACTAGTGGTGTCGGTCATCAA |
| Cassette8_3 | AATTGCTACCACCCCTTGCTGACCTACNNKTTTCAGCCCACTAGTGGTGTCGGTCATCAA |
| Cassette8_4 | AATTGCTACCACCCCTTGCTGACATATAACNNKCAGCCCACTAGTGGTGTCGGTCATCAA |
| Cassette8_5 | AATTGCTACCACCCCTTGCTGACATACAATTTCNNKCCCACTAGTGGTGTCGGTCATCAA |
| Cassette8_6 | AATTGCTACCACCCCTTGCTGACCTATAATTTCCAGNNKACTAGTGGTGTCGGTCATCAA |
| Cassette8_7 | AATTGCTACCACCCCTTGCTGACCTATAACTTTCAACCTNNKAGTGGTGTCGGTCATCAA |
| Cassette8_8 | AATTGCTACCACCCCTTGCTGACCTACAACTTCCAGCCTACTNNKGGTGTCGGTCATCAA |
| Cassette9_1 | TACAACTTTCAACCCACTAGTNNKGTAGGGCATCAACCGCATAGAGTGGTGGTTCTC |
| Cassette9_2 | TACAACTTTCAACCCACTAGTGGGNNKGGTCACCAACCGCATAGAGTGGTGGTTCTC |
| Cassette9_3 | TACAACTTTCAACCCACTAGTGGTGTANNKCACCAGCCGCATAGAGTGGTGGTTCTC |
| Cassette9_4 | TACAACTTTCAACCCACTAGTGGGGTCGGGNNKCAGCCGCATAGAGTGGTGGTTCTC |
| Cassette9_5 | TACAACTTTCAACCCACTAGTGGTGTAGGTCATNNKCCTCATAGAGTGGTGGTTCTC |
| Cassette9_6 | TACAACTTTCAACCCACTAGTGGTGTCGGGCACCAANNKCATAGAGTGGTGGTTCTC |
| Cassette9_7 | TACAACTTTCAACCCACTAGTGGGGTAGGTCATCAGCCGNNKAGAGTGGTGGTTCTC |

**Extended Data Table 3. Sequences of the reverse primers for constructing NJ-RBM mutant library.**

| Primer ID | Sequence (5' to 3') |
|---|---|
| Cassette1_R | CCATGCAATGACACAACCGGT |
| Cassette2_R | ATTGCTGTCAAGCGTTTTACT |
| Cassette3_R | CTTGTACTTGGTATTGCCCTT |
| Cassette4_R | GCGGCTCTTCCTCACGAAACG |
| Cassette5_R | GATGTCCCTTTCAAAGGGCTG |
| Cassette6_R | ACCCGCTTGAAAAATTTCAGT |
| Cassette7_R | GCCCACGGTATTGCATGGGCG |
| Cassette8_R | CAGCAAGGGGTGGTAGCAATT |
| Cassette9_R | ACTAGTGGGTTGAAAGTTGTA |

```
                      334 346 367 369 372 384
NJ-RBD                TKLCPFGEVFNATNFASVYAWNRKRISNCVADYSFLSNS-SFSTFKCYGVSHTKLNDLCF
WT-RBD                TNLCPFGEVFNATRFASVYAWNRKRISNCVADYSVLYNSASFSTFKCYGVSPTKLNDLCF
Alpha-RBD             TNLCPFGEVFNATRFASVYAWNRKRISNCVADYSVLYNSASFSTFKCYGVSPTKLNDLCF
Beta-RBD              TNLCPFGEVFNATRFASVYAWNRKRISNCVADYSVLYNSASFSTFKCYGVSPTKLNDLCF
Delta-RBD             TNLCPFGEVFNATRFASVYAWNRKRISNCVADYSVLYNSASFSTFKCYGVSPTKLNDLCF
Omicron_BA.1-RBD      TNLCPFDEVFNATKFASVYAWNRKRISNCVADYSVLYNLAPFFTFKCYGVSPTKLNDLCF
Omicron_BA.2-RBD      TNLCPFDEVFNATRFASVYAWNRKRISNCVADYSVLYNFAPFFAFKCYGVSPTKLNDLCF
Omicron_BA.4/5-RBD    TNLCPFDEVFNATRFASVYAWNRKRISNCVADYSVLYNFAPFFAFKCYGVSPTKLNDLCF
Omicron_BQ.1.1-RBD    TNLCPFDEVFNATTFASVYAWNRKRISNCVADYSVLYNFAPFFAFKCYGVSPTKLNDLCF
Omicron_XBB.1.5-RBD   TNLCPFHEVFNATTFASVYAWNRKRISNCVADYSVIYNFAPFFAFKCYGVSPTKLNDLCF
Omicron_BA.2.86-RBD   TNLCPFHEVFNATRFASVYAWNRTRISNCVADYSVLYNFAPFFAFKCYGVSPTKLNDLCF
Omicron_JN.1-RBD      TNLCPFHEVFNATRFASVYAWNRTRISNCVADYSVLYNFAPFFAFKCYGVSPTKLNDLCF
Omicron_KP.2-RBD      TNLCPFHEVFNATTFASVYAWNRTRISNCVADYSVLYNFAPFFAFKCYGVSPTKLNDLCF
Omicron_KP.3-RBD      TNLCPFHEVFNATRFASVYAWNRTRISNCVADYSVLYNFAPFFAFKCYGVSPTKLNDLCF
                      *:**** ****** *********.**********.: *   * :******* ********
```

```
                      403 408 413 415 421 439 440 444 445 446 449 450 452
NJ-RBD                TNVYADSFVIKGDEVTQIAPRQEGKIADFNYKLPDDFTGCVIAWNSKTLDSNNKGNTKYK
WT-RBD                TNVYADSFVIRGDEVRQIAPGQTGKIADYNYKLPDDFTGCVIAWNSNNLDSKVGGNYNYL
Alpha-RBD             TNVYADSFVIRGDEVRQIAPGQTGKIADYNYKLPDDFTGCVIAWNSNNLDSKVGGNYNYL
Beta-RBD              TNVYADSFVIRGDEVRQIAPGQTGNIADYNYKLPDDFTGCVIAWNSNNLDSKVGGNYNYL
Delta-RBD             TNVYADSFVIRGDEVRQIAPGQTGKIADYNYKLPDDFTGCVIAWNSNNLDSKVGGNYNYR
Omicron_BA.1-RBD      TNVYADSFVIRGDEVRQIAPGQTGNIADYNYKLPDDFTGCVIAWNSNKLDSKVSGNYNYL
Omicron_BA.2-RBD      TNVYADSFVIRGNEVSQIAPGQTGNIADYNYKLPDDFTGCVIAWNSNKLDSKVGGNYNYL
Omicron_BA.4/5-RBD    TNVYADSFVIRGNEVSQIAPGQTGNIADYNYKLPDDFTGCVIAWNSNKLDSKVGGNYNYR
Omicron_BQ.1.1-RBD    TNVYADSFVIRGNEVSQIAPGQTGNIADYNYKLPDDFTGCVIAWNSNKLDSTVGGNYNYR
Omicron_XBB.1.5-RBD   TNVYADSFVIRGNEVSQIAPGQTGNIADYNYKLPDDFTGCVIAWNSNKLDSKPSGNYNYL
Omicron_BA.2.86-RBD   TNVYADSFVIKGNEVSQIAPGQTGNIADYNYKLPDDFTGCVIAWNSNKLDSKHSGNYDYW
Omicron_JN.1-RBD      TNVYADSFVIKGNEVSQIAPGQTGNIADYNYKLPDDFTGCVIAWNSNKLDSKHSGNYDYW
Omicron_KP.2-RBD      TNVYADSFVIKGNEVSQIAPGQTGNIADYNYKLPDDFTGCVIAWNSNKLDSKHSGNYDYW
Omicron_KP.3-RBD      TNVYADSFVIKGNEVSQIAPGQTGNIADYNYKLPDDFTGCVIAWNSNKLDSKHSGNYDYW
                      **********:*:** **** * *:***:*****************:.***.  ** .*
```

```
                      453 455 456 460 462 473 477 478 482 483 484 486 490 493 496 501 505 508
NJ-RBD                FRFVRKSRLQPFERDISTEIFQAGNRPCNT-VGLNCYHPLLTYNFQPTSGVGHQPHRVVV
WT-RBD                YRLFRKSNLKPFERDISTEIYQAGSTPCNGVEGFNCYFPLQSYGFQPTNGVGYQPYRVVV
Alpha-RBD             YRLFRKSNLKPFERDISTEIYQAGSTPCNGVEGFNCYFPLQSYGFQPTYGVGYQPYRVVV
Beta-RBD              YRLFRKSNLKPFERDISTEIYQAGSTPCNGVKGFNCYFPLQSYGFQPTYGVGYQPYRVVV
Delta-RBD             YRLFRKSNLKPFERDISTEIYQAGSKPCNGVEGFNCYFPLQSYGFQPTNGVGYQPYRVVV
Omicron_BA.1-RBD      YRLFRKSNLKPFERDISTEIYQAGNKPCNGVAGFNCYFPLRSYSFRPTYGVGHQPYRVVV
Omicron_BA.2-RBD      YRLFRKSNLKPFERDISTEIYQAGNKPCNGVAGFNCYFPLRSYGFRPTYGVGHQPYRVVV
Omicron_BA.4/5-RBD    YRLFRKSNLKPFERDISTEIYQAGNKPCNGVAGVNCYFPLQSYGFRPTYGVGHQPYRVVV
Omicron_BQ.1.1-RBD    YRLFRKSKLKPFERDISTEIYQAGNKPCNGVAGVNCYFPLQSYGFRPTYGVGHQPYRVVV
Omicron_XBB.1.5-RBD   YRLFRKSKLKPFERDISTEIYQAGNKPCNGVAGPNCYSPLQSYGFRPTYGVGHQPYRVVV
Omicron_BA.2.86-RBD   YRLFRKSKLKPFERDISTEIYQAGNKPCKG-KGPNCYFPLQSYGFRPTYGVGHQPYRVVV
Omicron_JN.1-RBD      YRSFRKSKLKPFERDISTEIYQAGNKPCKG-KGPNCYFPLQSYGFRPTYGVGHQPYRVVV
Omicron_KP.2-RBD      YRSLRKSKLKPFERDISTEIYQAGNKPCKG-KGPNCYFPLQSYGFRPTYGVGHQPYRVVV
Omicron_KP.3-RBD      YRSLRKSKLKPFERDISTEIYQAGNKPCKG-KGPNCYFPLESYGFRPTYGVGHQPYRVVV
                      :* .***.*:**********:***. **:   * *** ** :*.*:** ***:**:****
```

```
                      519 529
NJ-RBD                LSFELLQAPATVCGPKRS
WT-RBD                LSFELLHAPATVCGPKKS
Alpha-RBD             LSFELLHAPATVCGPKKS
Beta-RBD              LSFELLHAPATVCGPKKS
Delta-RBD             LSFELLHAPATVCGPKKS
Omicron_BA.1-RBD      LSFELLHAPATVCGPKKS
Omicron_BA.2-RBD      LSFELLHAPATVCGPKKS
Omicron_BA.4/5-RBD    LSFELLHAPATVCGPKKS
Omicron_BQ.1.1-RBD    LSFELLHAPATVCGPKKS
Omicron_XBB.1.5-RBD   LSFELLHAPATVCGPKKS
Omicron_BA.2.86-RBD   LSFELLHAPATVCGPKKS
Omicron_JN.1-RBD      LSFELLHAPATVCGPKKS
Omicron_KP.2-RBD      LSFELLHAPATVCGPKKS
Omicron_KP.3-RBD      LSFELLHAPATVCGPKKS
                      ******:*********:*
```

**Extended Data Fig 1. Sequence alignment of RBDs including cryptic lineage NJ and several VOC/VOIs.** Red indicates unique substitutions in NJ-RBD. Blue indicates convergent substitutions between NJ-RBD and other VOC/VOI RBDs. Residues identical in all aligned sequences are labeled by an asterisk (*), whereas a colon (:) and a period (.) indicate highly similar and less similar sequences, respectively. The sequence alignment was performed using Clustal Omega[2].

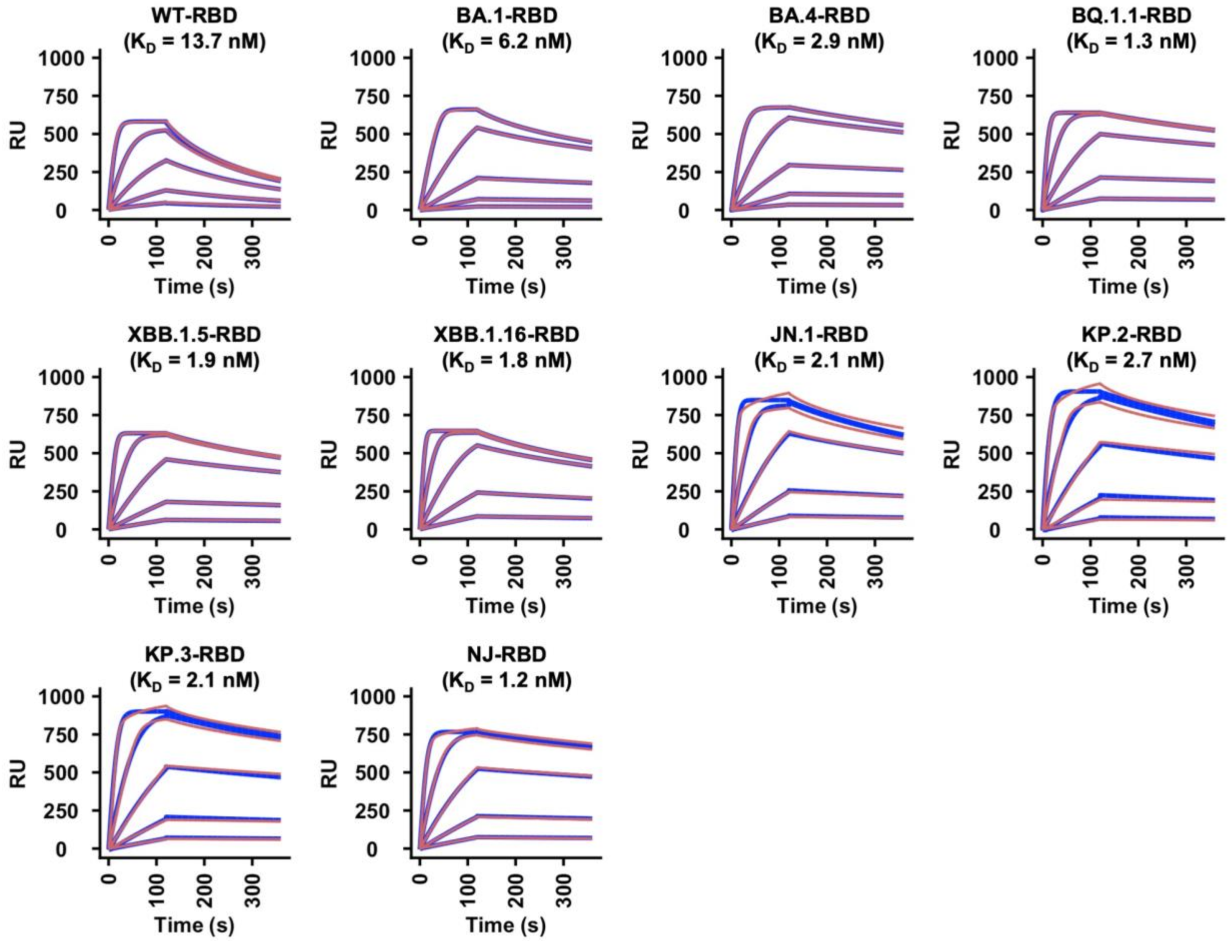


**Extended Data Fig 2. Receptor binding of various RBDs to hACE2 using SPR.** Biacore sensorgram representing the binding of the indicated RBD to hACE2. The y-axis shows the response unit (RU). Red lines represent the response curve, and blue lines represent a 1:1 binding model. Binding kinetics were measured against hACE2 at 0 nM, 3.7 nM, 11.1 nM, 33.3 nM, 100 nM, and 300 nM. Dissociation constant ($K_D$) is indicated.

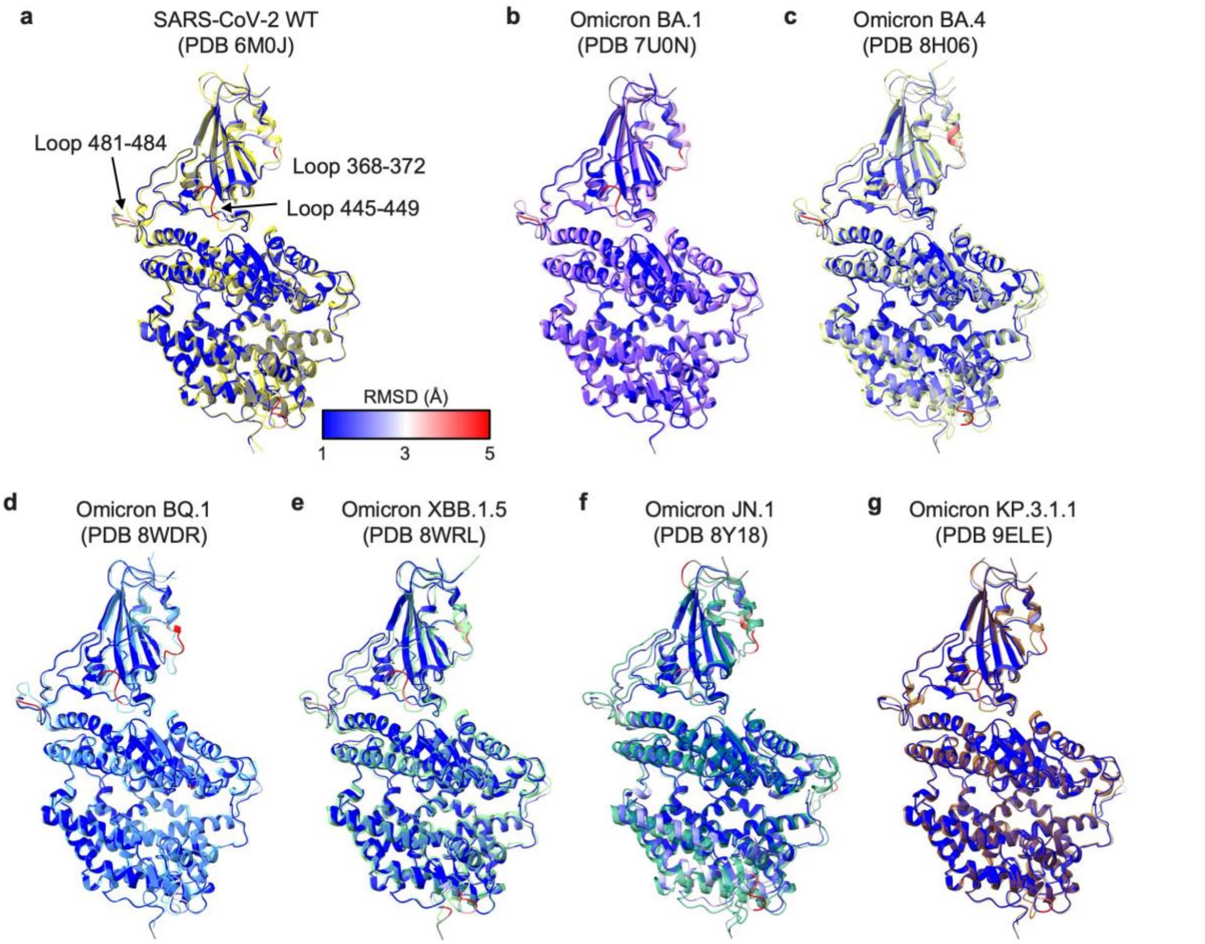


**Extended Data Fig 3. Structural comparison between hACE2 in complex with NJ-RBD and in complex with RBDs from other VOCs/VOIs.** Positions of loops 368-372, 445-449, and 481-484 are labeled. Structural alignment of the NJ-RBD/hACE2 structure (this study, blue) to hACE2 in complex with **(a)** SARS-CoV-2 WT (PDB 6M0J, yellow)[3], **(b)** Omicron BA.1 (PDB 7U0N, pink)[4], **(c)** BA.4 (PDB 8H06, lime)[5], **(d)** BQ.1 (PDB 8WDR, cyan)[6], **(e)** XBB.1.5 (PDB 8WRL, light green)[7], **(f)** JN.1 (PDB 8Y18, green)[8], and **(g)** KP.3.1.1 (PDB 9ELE, brown)[9], respectively. Structural differences of NJ-RBD/hACE2 are color-coded by their root mean square deviation (RMSD) (Å).

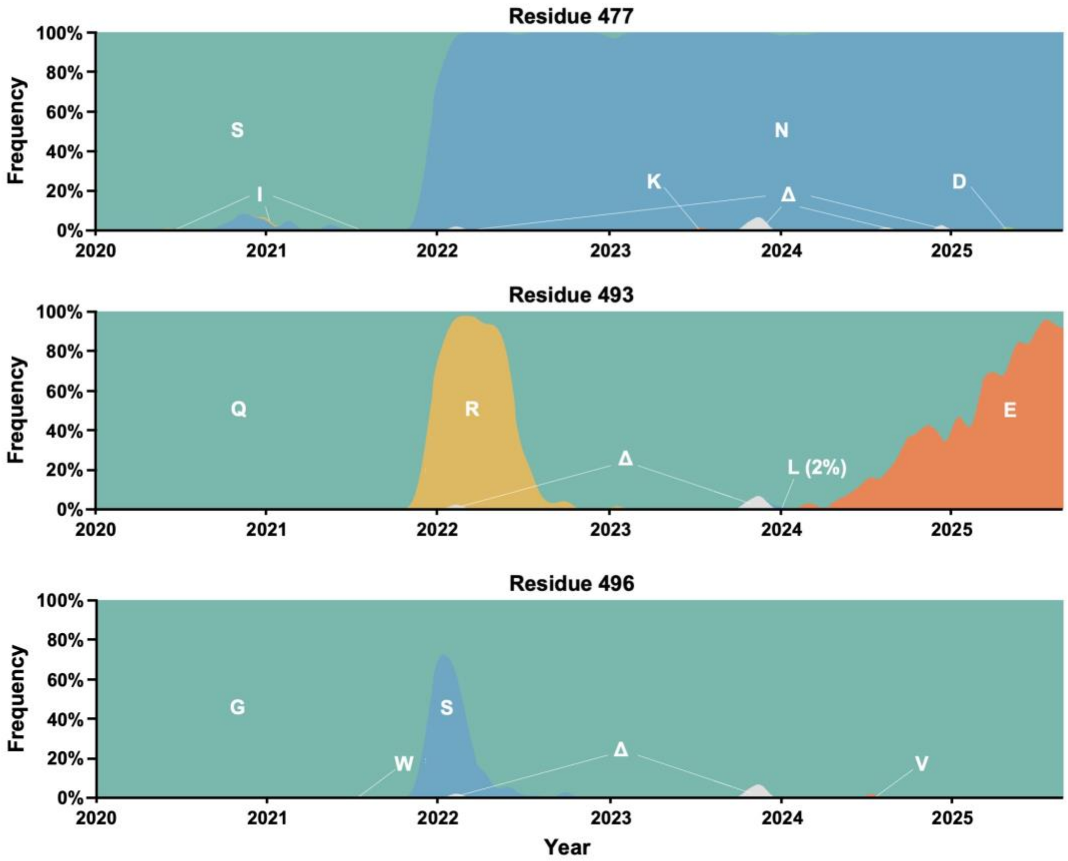


**Extended Data Fig 4. Frequency of amino acids at positions 477, 493, and 496 of SARS-CoV-2 spike as of July 2025.** Plot was generated using Nextstrain (nextstrain.org)[10], which sources data from GISAID[11]. "Δ" indicates deletion.

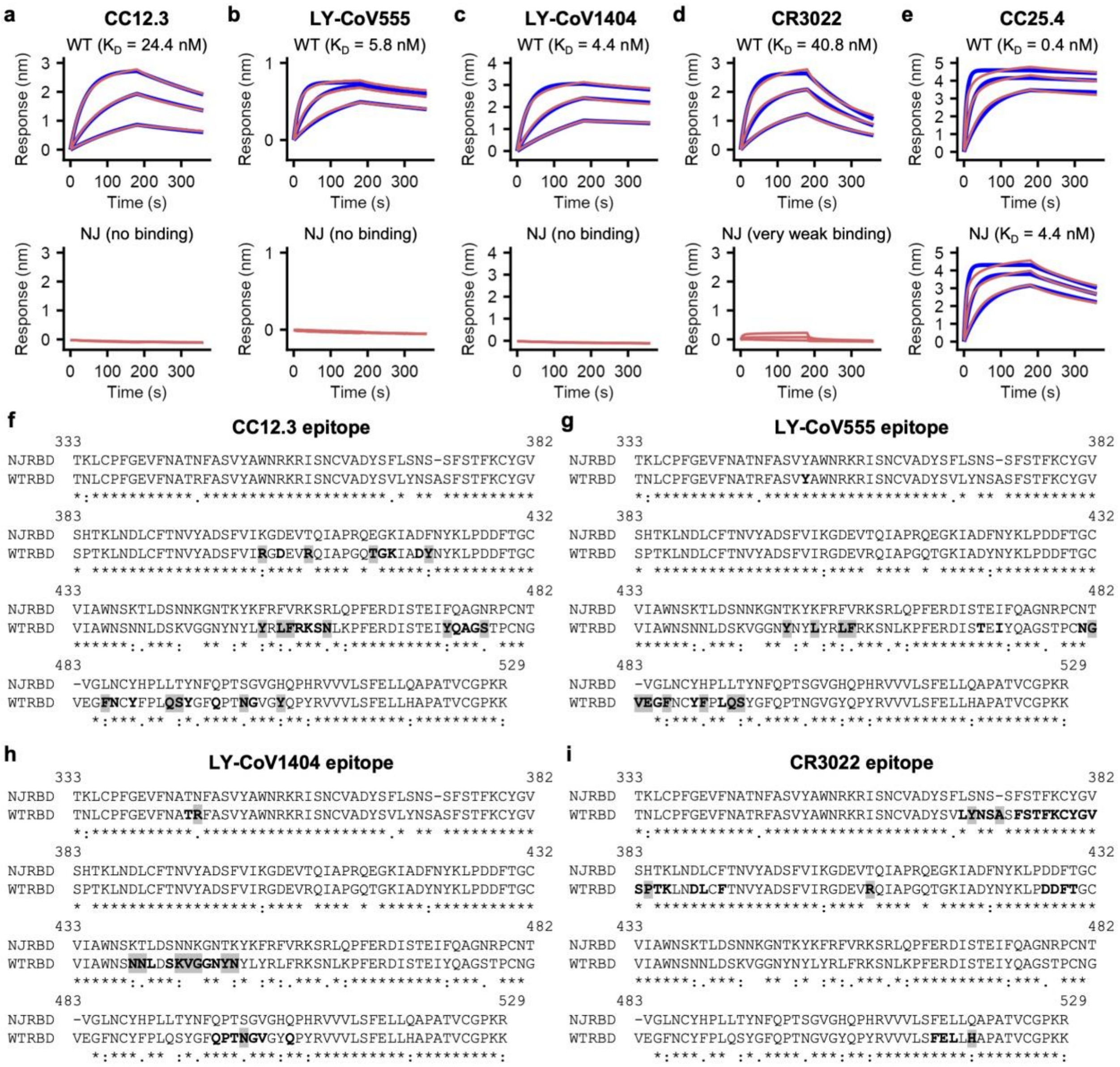


**Extended Data Fig 5. Sensorgrams for binding of WT-RBD and NJ-RBD to neutralizing antibodies.** Binding kinetics of WT-RBD and NJ-RBD against **(a)** CC12.3, **(b)** LY-CoV555, **(c)** LY-CoV1404, **(d)** CR3022, and **(e)** CC25.4 Fabs were measured by biolayer interferometry (BLI). Red lines represent the response curve. Blue lines represent a 1:1 binding model. Binding kinetics were measured for antibody concentrations at 33.3 nM, 100 nM, and 300 nM. Dissociation constant ($K_D$) is indicated. **(f-i)** Sequence alignment of NJ-RBD and WT-RBD with the indicated epitope residues (BSA > 0 Å$^2$ as calculated by PISA[12]) in bold. Non-conserved epitope residues are highlighted in grey.

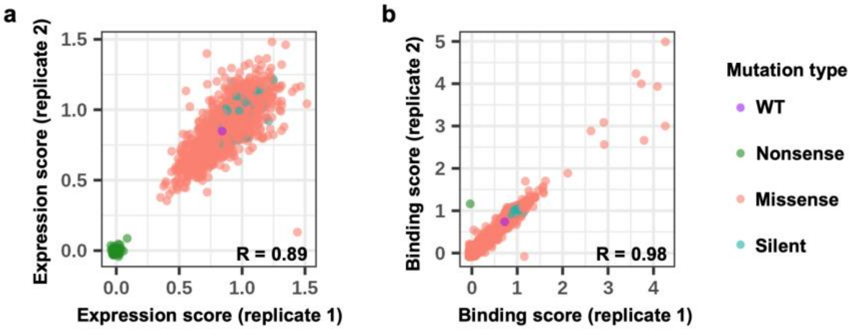


**Extended Data Fig 6. Correlation between biological replicates of binding and expression sorts of NJ-RBM variant library. a**, Correlation of expression scores between two independent biological replicates is shown. **b**, Correlation of binding scores between two independent biological replicates is shown. Mutation types are color-coded as follows: pink (missense), green (nonsense), blue (silent), and purple (WT). The Pearson correlation coefficient (R) is indicated. Please see Figure 5 for expression and binding analyses.
